# Real-Time HIIT Response in Patients with Coronary Artery Disease: A CERT-Based Report from the Heart-Brain Randomized Controlled Trial

**DOI:** 10.1101/2025.07.23.25332034

**Authors:** Javier Fernández-Ortega, Angel Toval, Lucía Sánchez-Aranda, Patricio Solis-Urra, Carlos Prieto, Rosa María Alonso-Cuenca, Alberto González-García, Esmée A. Bakker, Isabel Martín-Fuentes, Beatriz Fernandez-Gamez, Marcos Olvera-Rojas, Andrea Coca-Pulido, Darío Bellón, Alessandro Sclafani, Javier Sanchez-Martinez, Ricardo Rivera-López, Norberto Herrera-Gómez, Rafael Peñafiel-Burkhardt, Víctor Manuel López-Espinosa, Sara Corpas-Pérez, Emilio J. Barranco-Moreno, Francisco J. Morales-Navarro, Raúl Nieves, Alfredo Caro-Rus, Francisco J. Amaro-Gahete, Sol Vidal-Almela, Anna Carlén, Dorthe Stevensold, Jenna L. Taylor, Eduardo Moreno-Escobar, Rocío García-Orta, Irene Esteban-Cornejo, Francisco B. Ortega

## Abstract

**Aims:** High-intensity interval training (HIIT) and resistance training (RT) are promising options for cardiac rehabilitation, yet exercise trials seldom report sufficient protocol details or participants’ response and experience in a standardised way. We therefore quantified and compared attendance rate, heart-rate–based compliance, perceived exertion, enjoyment and affect during a 12-week programme performed as either HIIT or HIIT plus RT (HIIT + RT). Both interventions were documented according to the Consensus on Exercise Reporting Template (CERT).

**Methods:** A total of 105 patients with coronary artery disease (62 ± 7 years, 78 % men) were randomised to: usual care, HIIT or HIIT + RT. This study focus on the two exercise programs (n=64) consisting on three supervised 45-min sessions per week. Intensity for the high-intensity intervals was prescribed as 85-95 % of peak heart rate (HRpeak) and monitored both objectively in real time (second-by-second) and subjectively (Borg 0–10 Rating of Perceived Exertion scale). Outcomes were (i) attendance (sessions attended / sessions offered), (ii) intensity compliance (proportion of sessions meeting intensity target), (iii) enjoyment (Physical Activity Enjoyment Scale, 1–7) and (iv) affect response (Feeling Scale, –5 to +5 pre- and post-exercise).

**Results:** Participants attended 88 % of scheduled sessions (HIIT 85 %, HIIT + RT 90 %; *p*=0.46) and were compliant with the prescribed heart rate intensity zone in an average of 75% of the attended sessions (HIIT 72%, HIIT+RT 78%; p = 0.48). Enjoyment was similarly high in both groups (overall PACES 5.4 / 7). Mean RPE during high-intensity intervals was 7.1 / 10 for both groups; in HIIT + RT, average RPE during resistance circuits were 5.1. Feeling-Scale scores improved after exercise sessions in both programs (HIIT +0.65; HIIT + RT +0.45), with a statistically significant advantage for HIIT+RT in post-session affective response, after adjusting for pre-session scores (p < .001)

**Conclusion:** Our HIIT and HIIT + RT programs resulted in high attendance and compliance, and were positively experienced by patients with coronary artery disease, providing a feasible and time-efficient alternative for meeting international exercise guidelines (including RT). The fully CERT-documented protocols offer a reproducible research and clinical work.

## Introduction

Coronary artery disease (CAD) is one of the leading causes of global mortality and morbidity in adults over 50 years of age (1,2), which consequently implies a high economic burden (2). CAD impairs heart function by restricting blood flow in the coronary arteries. Over time, it can lead to angina or myocardial infarction (3). Moreover, CAD is associated with many other comorbidities (4), including an increased risk of cognitive decline in comparison with healthy peers (2). Although pharmacological treatments have long represented the cornerstone of CAD management, current clinical guidelines advocate for a comprehensive approach that combines both pharmacological and non-pharmacological strategies (3). Among the latter, physical exercise has emerged as a highly effective intervention to improve cardiometabolic health in patients with CAD (5), as well as improving their quality of life and mental health (6).

Despite the well-known health-related benefits of exercise, exercise-based cardiac rehabilitation (CR) programs remain underutilised, partly due to low attendance rates, which represent a major challenge in maximizing their impact (7,8). Previous evidence highlight several factors that potentially explain this issue, including (i) patient-related barriers (e.g., older age, female sex, lower education, multiple comorbidities, lack of social support, and competing family responsibilities), and (ii) logistical constraints (e.g., transportation difficulties and distance from facilities) (9–11). Furthermore, sustaining physical activity levels after completing CR remains a significant challenge, with key determinants of long-term adherence including self-motivation, enjoyment, and self-efficacy (12). Specifically, enjoyment levels mediate the relationship between exercise motivation and sustained physical activity (13,14). Besides attendance and enjoyment, compliance with the exercise prescription during the exercise sessions is also essential. Accurate regulation and monitoring of exercise intensity are essential to avoid overexertion, abnormal clinical signs, or injury, and also to ensure that patients receive an adequate physiological stimulus, especially in high-intensity protocols, to elicit meaningful cardiovascular and metabolic adaptations (15–19). Due to its complexity, regulating exercise intensity often demands the greatest involvment from cardiac rehabilitation professionals (20).

Historically, aerobic training, particularly moderate-intensity continuous training (MICT), has been the most widely studied modality for CAD management (21). Recent evidence suggests comparable or superior effects of high-intensity interval training (HIIT) *versus* MICT for improving cardiorespiratory fitness in CR patients (22), and particularly in those with CAD (23–26). Notably, the influence of resistance training remains underexplored in CAD populations, even though it is recommended by the World Health Organization (WHO) physical activity guidelines and by CAD clinical management guidelines (3,27). These guidelines advocate for a combined approach integrating aerobic and resistance exercise, which appears particularly promising for improving functional capacity in individuals with CAD (15).

Despite the growing literature evaluating the effects of exercise on health, most exercise interventions are poorly reported. Key aspects, such as type and dose of exercise, supervision, attendance and compliance rates, enjoyment, or contextual factors, are often insufficiently described, limiting both reproducibility and clinical translation (21). This is particularly problematic in CAD, where interventions tend to be complex and highly individualized, making adequate reporting essential for evaluating feasibility and efficacy across settings (28). High-quality reporting of exercise interventions requires precise description of the exercise protocol, supervision and monitoring procedures, adherence metrics, participants’ subjective experience, safety outcomes, and the broader intervention context. Such comprehensive reporting is essential not only to enhance the reproducibility and interpretation of trial findings, but also to ensure reliable results and support the translation of evidence into clinical practice. Clear and rigorous descriptions of these components enable clinicians and policymakers to evaluate the effectiveness and feasibility of interventions, and to implement them confidently in real-world settings (21,28). For that purpose, the Consensus on Exercise Reporting Template (CERT) was developed to standardize the reporting of exercise interventions in clinical trials. The CERT is a 16-item checklist designed to structure the information, promote transparency, improve results interpretation and replication, and facilitate an effective implementation into practice of the trial interventions (29,30). Well-designed CERT-based randomised controlled trials (RCTs) should accurately report detailed information about the FITT-VP exercise principles (i.e., Frequency, Intensity, Time, Type, Volume and Progression) to establish accessible, reproducible and robust evidence-based exercise recommendations for enhancing physical and brain health in both healthy and diseased individuals, considering the unique characteristics of the population under study.

The aims of this study are two-fold. First, to provide a comprehensive CERT-based description and rationale for two supervised exercise interventions implemented in the Heart-Brain RCT in patients with CAD: (1) HIIT, and (2) HIIT+RT. Second, to quantify and compare these interventions in terms of session attendance, intensity compliance assessed via second-by-second heart rate monitoring and perceived exertion, as well as affective responses and enjoyment. These outcomes aim to inform the optimization of exercise prescriptions in future interventions targeting cognitive and brain health outcomes in individuals with CAD

## Methods/design

### Study design

Detailed information about the study protocol of the Heart-Brain RCT has been published elsewhere (31). Briefly, a total of 105 adults with a diagnosis with CAD (aged 50-75 years old) from Granada (Spain) were randomised on a rolling basis after completing baseline assessments, using a 1:1:1 ratio and stratified by age (<65 or ≥65 years) and sex. Allocation was concealed and carried out by a blinded external researcher into three groups: 1) usual care control group (n=33), 2) HIIT (n=35), and 3) HIIT+RT(n=37). The present study only focuses on the two exercise groups, which underwent 12-week supervised exercise interventions aiming to evaluate their effects on cerebral blood flow as the primary outcome, and different brain and cardiovascular health parameters as secondary outcomes.

Eligible patients received written information and provided informed consent before participation. The study is registered at ClinicalTrials.gov (Identifier: NCT06214624). The trial was approved by the Research Ethics Board of the Andalusian Health Service (CEIM/CEI Provincial de Granada; #1776-N-21on December 21st, 2021)

### Exercise program rationale

The rationale of the two exercise interventions implemented in the Heart-Brain project was described following the CERT criteria recommendations shown in Supplementary Material Table 1, and the structure and characteristics of the interventions are summarised in Figure 1.

**Fig. 1.**
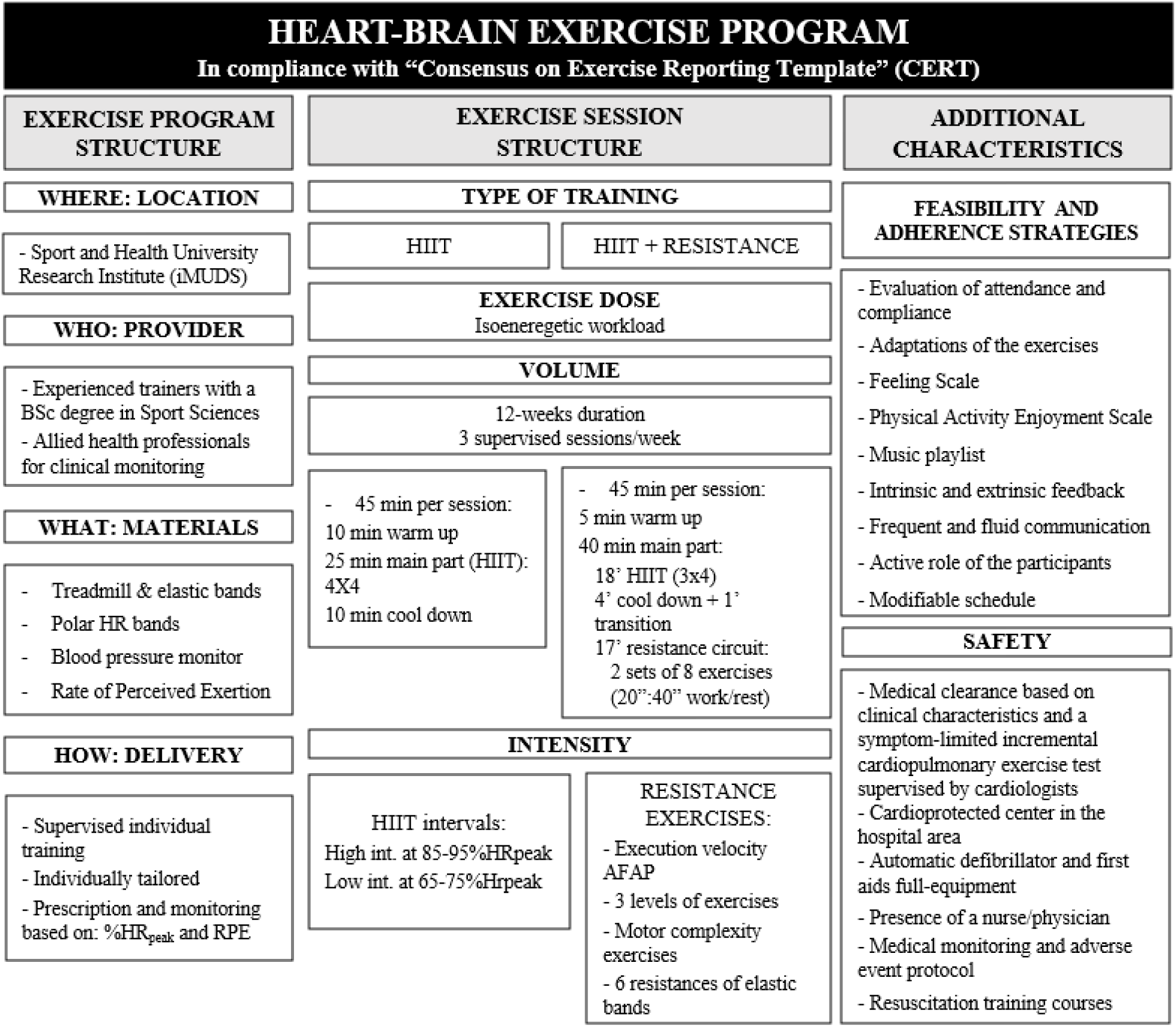
Graphical abstract of the exercise interventionsimplemented in the Heart-Brain study. HIIT: High Intensity Interval Training; AFAP: As fast as possible; HR: Heart rate; RPE: Rate of perceived exertion.

Patients were allocated to one of two exercise interventions with distinct session structures as shown in Figure 2. One intervention consists of aerobic high-intensity interval training, HIIT, while the other involves concurrent training, combining aerobic HIIT and resistance training, HIIT+RT. Both interventions consisted of 12 weeks of supervised exercise, with 3 sessions per week and a duration of 45 minutes per session. The initial two weeks are dedicated to familiarization, allowing patients to progressively adapt to this type of training and to determine the appropriate speed and incline required to reach their individually prescribed intensity zones. The HIIT modality was designed based on the guidelines for the delivery and monitoring of HIIT in clinical populations (32). Additionally, both interventions align with the physical activity WHO guidelines for adults, as it exceeds the minimum requirement of 75 minutes of vigorous-intensity or 150 minutes of moderate-intensity exercise per week (33). Furthermore, the HIIT+RT intervention also meets WHO guidelines for resistance training involving large muscle groups, as it surpasses the required two sessions per week. The HIIT+RT program was designed to meet all the recommendations in a time efficient manner (i.e., 45min x 3 times/week).

**Fig. 2.**
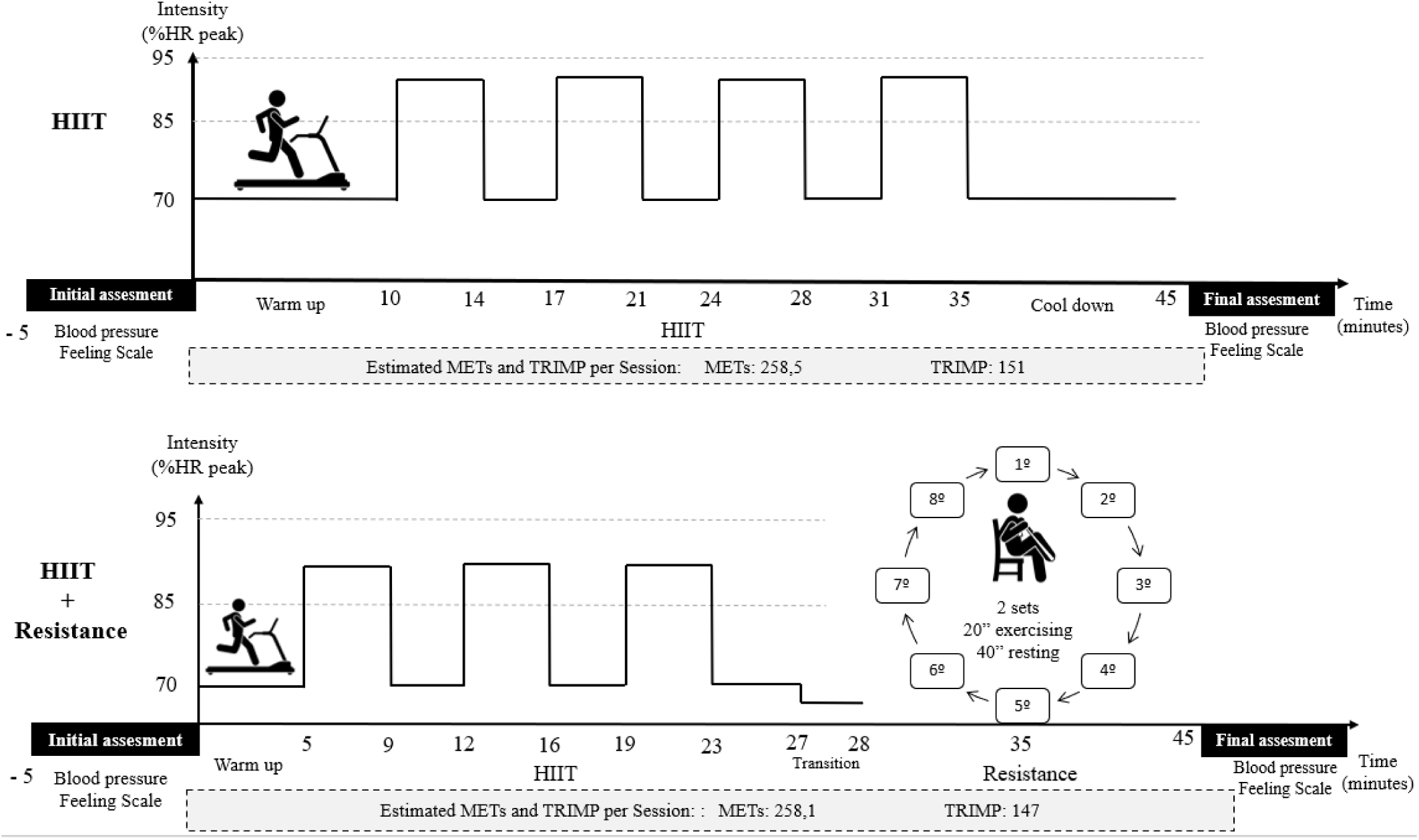
Details of the session structure of the HIIT-based (above) and the HIIT + RT (below) exercise interventions, (**HIIT:** high-intensity interval training; **MET:** metabolic equivalent of task; **TRIMP:** training impulse (session load).

#### Description of HIIT-based exercise intervention

The HIIT training was designed to be performed in a treadmill. The warm-up consisted of 10 minutes of walking with gradual increase of the speed and/or slope, to reach and keep 70% heart rate peak (HR_peak_) at least during the last 5 minutes (Figure 2). In the last 30 seconds of the warm-up the patient was asked about the rating of perceived exertion (RPE), which was recorded together with the %HRpeak, speed and slope. The recording of these parameters was repeated after each high-intensity interval. The main phase had a total time of 25 minutes and consisted of 4-minute x 4 intervals of high-intensity. During these intervals, the target was to reach 85-95% of HRpeak and maintain it throughout the entire interval. In the first interval, the target intensity should be reached at least during the last 30 seconds, and in the next three intervals, the target intensity must be reached and maintained during the entire second half of the interval (32). Each high-intensity interval was followed by a 3 minute recovery period, during which the goal was to maintain ∼70% of HRpeak to optimize recovery while sustaining cardiovascular engagement. The %HRpeak was increased by adjusting the speed and/or slope, selecting the option that was most feasible for each patient. In most cases, patients walked with varying inclines throughout the HIIT training. However, in cases of pain or inability to tolerate slope increases, patients performed a fast walk or even ran instead (those able to do so). The cool-down phase lasts 10 minutes where the speed and slope were gradually decreased to lower the HR.

The *HIIT+RT group* performed the *HIIT* training first, and secondly RT, as shown in Figure 2. The HIIT comprised a warm-up, a main part and a cool-down phase. The warm-up consisted of 5 minutes of a gradual increase of the intensity to reach the 70% of HRpeak. The main phase had a duration of 18 minutes: 4-minute x 3 intervals at high-

intensity 85-95% of HRpeak, and 2 recovery intervals of 3 minutes in between at lower intensity ∼70% of HRpeak. Monitorization and progression of the intensity was similar to the HIIT-based intervention described above. The cool-down lasts 4 minutes where the speed and slope were gradually decreased to reduce the HR. A transition period of 1 minute and 20 seconds was included between the treadmill and resistance exercises to prevent any dizziness caused by the treadmill. The RT part consists of a circuit of 8 exercises with 2 sets. Each exercise was performed for 20 seconds, followed by 40 seconds of rest (which included the transition between workout stations). Additionally, there was an additional one-minute rest period between the first and second circuit (detailed description of the exercises in Supplementary Material Table 2). The exercises were organized in a specific sequence that alternates major movement patterns, including vertical and horizontal pushes and pulls, lower body movements, and core stabilization, as detailed in Supplementary Material Table 3. In general, each session included one exercise per major muscle group, aligned with different movement patterns (e.g., vertical push, core, legs). In some cases, two exercises were available for the same muscle group, and these were alternated across sessions to introduce variety and avoid overuse.

### Location, providers, and materials (where, who, and what)

The interventions were carried out at the Sport and Health University Research Institute (iMUDS), which hosts the Andalusian Center of Sports Medicine (CAMD), located by the Hospital area (200m apart) in the Technological Health Park of Granada (Spain). The center was fully equipped to perform aerobic and resistance training. Detailed description of the exercise equipment is shown in Supplementary Material Table 4. In the resistance exercise part of the intervention, patients performed body-weight exercises and used elastic bands (Thera-Band®) for added resistance. This exercise approach was partially implemented in one of our previous trials, demonstrating its feasibility in similar clinical populations (older adults) (34,35) These bands are available in 6 different colors corresponding to different resistances (i.e., yellow-soft, red-medium, green-strong, blue-extra strong, black-strong special, silver-athletic, and gold-olympic) and have been tested in previous studies to ensure consistent tension levels (36,37). All the elastic bands have a length of 1.50 m and were tied to fixed structures.

### Delivery of the training (how)

All sessions were supervised and delivered individually, with a 1:1 trainer-to-patient ratio. In rare cases, a 1:2 ratio was applied due to patient-driven scheduling changes,.

All participants underwent a cardiopulmonary exercise test (CPET) with ECG monitoring while taking their regular cardiovascular medications (including β-blockers), thereby reflecting their habitual physiological status at baseline. HRpeak was calculated using a hierarchical approach: when available, it was defined as the highest 5-second average heart rate recorded by the Polar device during the final 30 seconds of exercise. If Polar data were unavailable, the highest HR recorded during the final ECG stage (1–30 seconds, depending on when the test ended) was used. If the penultimate ECG stage showed a higher value than the final one, that value was selected as HRpeak.

Exercise prescription, monitoring, and progression decisions for the HIIT component were individualized based on the percentage of HRpeak and RPE on a 0–10 scale (38). In contrast, during the resistance training component of the HIIT+RT intervention, progression was guided by the trainer’s clinical judgment, as RPE was not considered a reliable indicator in this context. This was primarily due to participants’ limited experience with resistance training, often reporting low RPE values (e.g., 3/10) even when reaching muscular failure. Additionally, blood pressure and symptoms were systematically monitored before and after each session. In-session monitoring was performed only in specific cases where it was clinically indicated for patient safety. Patients taking beta-blocker medications were expected to display lower absolute heart rate values during exercise sessions compared to those not on beta-blockers, even when exercising at a similar relative intensity, due to the chronotropic effects of the medication (32). Thus, while the HRpeak was used as a reference for exercise intensity prescription, patients were encouraged to use RPE (39–42) in cases of discordance between HR and RPE targets (32). In addition to HR and RPE, the judgement of the trainers was the last criteria for intensity adaptations, particularly in the RT, where RPE might be more difficult to rate by unexperienced patients.

### Exercise sessions characteristics: type and doses

#### General session structure

The structure of all exercise sessions were as follows: 1) initial assessment (first, patients sit for five minutes to measure the blood pressure in resting condition, then patients answered the Feeling Scale (43), 2) warm-up, 3) aerobic training, 4) RT (only for the HIIT+RT group), 5) cool-down, and 6) final assesments (blood pressure and Feeling Scale) (Figure 2).

#### Frequency, length of the session and program duration

The two intervention groups exercised three times per week, for 12 weeks (36 sessions in total). Each HIIT session lasted 45 minutes and each HIIT + RT lasted 45 minutes. Generally, the sessions were carried out on Monday, Wednesday and Friday to allow a sufficient recovery in between. Training sessions were scheduled either in the morning or afternoon, depending on each patient’s availability. Although patients typically trained at the same time of day throughout the intervention, occasional schedule adjustments were made to facilitate attendance. The maximum total training time for one patient was 27 hours (if completing the 36 sessions).

#### Intensity

HR was continuously recorded in every session with a Polar H10 chest-strap monitor (44,45). Five intensity zones, defined as percentage peak HR, were used to guide training (Supplementary Material Table 5). During the HIIT segments of both intervention arms, treadmill speed and/or incline were adjusted on-the-fly whenever necessary to keep each participant within the prescribed target zone and ensure a consistent stimulus across sessions.

In the RT, the exercises were performed with an emphasis on power (46), executing the concentric phase as fast as possible and the eccentric phase at a slower pace. Proper technique was consistently prioritised throughout the sessions. To adjust the intensity throughout the 12 weeks, three different levels of exercise were implemented, along with varying resistances. After each exercise, patients were asked to rate their effort using the RPE scale. Progression to the next level of difficulty and the use of stronger elastic bands were determined based on the patient’s RPE and the trainer’s assessment of proper exercise technique.

#### Total training load

Total training load was quantified with the TRaining IMPulse (TRIMP) method (47), and the session-by-session scores are presented in Supplementary Table 6. In this study we used a modified TRIMP, calculated as the product of effective training time (min) and the mean relative intensity, expressed as the percentage of each patient’s HRpeak obtained during the baseline cardiopulmonary exercise test.

#### Isoenergetic calculations

Both exercise interventions have been matched on energy expenditure to provide an isoenergetic workload in terms of intensity and duration, as shown in the isoenergetic calculations section and Supplementary Material Table 7. The isoenergetic calculations of the training interventions were based on the metabolic equivalents of tasks (MET) for exercise prescription defined by the compendium of the American College of Sports Medicine (19,48). For these calculations, 70% of HRpeak was equivalent to 4.5 MET, while 90% of HRpeak was equivalent to 8 MET. Resistance exercises were defined by the compendium as 5.3 MET (48).Therefore, the total MET per session for HIIT was 258.5 MET-min, calculated as 29 minutes at 4.5 MET plus 16 minutes at 8 MET. Over the course of a week, this resulted in 775.5 MET-min/week. The total MET per session for HIIT+RT was 258.1 MET-min, calculated as 16 minutes at 4.5 MET plus 12 minutes at 8 MET plus 17 minutes at 5.3 MET. Over the course of a week, this resulted in 774.3 MET-min/week. Therefore, the training volume for both groups was comparable and designed to achieve the 2020 WHO physical activity guidelines of 600 MET-min/week.

### Additional characteristics of the programs

#### Feasibility

The feasibility of Heart-Brain exercise interventions was recorded and evaluated by: the modifications and adaptations of exercises, the attendance, the intensity compliance, the affective response, the enjoyment and the occurrence of adverse events.

#### Safety and adverse events

The HIIT protocol used in this study was selected based on its proven safety profile in patients with CAD, having been previously implemented at a large scale and associated with low rates of serious adverse events in well-documented clinical settings (49).

The iMUDS is officially certified as a “Cardioprotected Center” by the Andalusian Health Authority (Decreto 22/2012, February 14th), and is fully equipped with an emergency trolley, including a first aid kit and an automated external defibrillator (AED). A trained healthcare professional (nurse or physician) was always present during the sessions to provide immediate assistance in the case of an adverse event. Furthermore, all members of the research and training team received specialized training in cardiopulmonary resuscitation (CPR) and defibrillator use.

To minimise risk, every participant first underwent a comprehensive medical evaluation to rule out absolute or relative contraindications to exercise. The screening included a cardiopulmonary exercise test, resting echocardiography, and a review of current medications and comorbidities to certify eligibility for the planned training loads. Once enrolled, patients were continuously overseen by cardiologists specialised in exercise prescription; blood pressure was recorded before every session and, when elevated, re-checked during exercise to guarantee real-time safety. In parallel, cardiac specialist nurses provided brief health-education touchpoints at each visit, emphasising the expected effects and potential adverse symptoms of prescribed medications, the acute responses to exercise, and general lifestyle recommendations to reinforce self-management outside the clinic.

The study adhered to the guidelines set forth by the Spanish Council of Cardiopulmonary Resuscitation (CERCP) for monitoring adverse events (50). Any adverse occurrences experienced by patients during the trial, including incidents of falls, musculoskeletal injuries, major cardiovascular events, or other health problems linked or not to the study’s procedures, were thoroughly documented, examined, and categorised by a cardiologist. Each event was documented individually within each study group.

Those in the exercise groups were given the opportunity to inform staff about any adverse events during their supervised sessions, which took place three times per week, whereas those in the control group were told to report incidents as soon as they happened. Moreover, patients in the control group were telephoned at weeks 3, 9, and 12 to track any potential adverse events.

An experienced cardiologist evaluated all events using the Liverpool Causality Assessment Tool (51) to ascertain whether each event was linked to the exercise intervention. Each adverse event was documented according to the classification and severity criteria outlined in the Common Terminology Criteria for Adverse Events (CTCAE) (52). The study’s cardiology team participated in collaborative discussions to classify events and conduct clinical assessments, which enabled accurate and consistent reporting to be carried out.

#### Modifications and adaptations of exercises

The HIIT phase was designed to be carried out on a treadmill. If a participant could not reach the prescribed intensity because of physical limitations, a pre-planned adaptation allowed the session to be completed on an alternative ergometer (cycle or cross-trainer). In the HIIT+RT group, in cases of injury, pain, or motor limitations, resistance exercises were either adapted to maintain the stimulus in the same muscle group with proper technique and safety, or, when adaptation was not feasible, the exercise was not performed.

### Monitoring of exercise response

#### Adherence: attendance and intensity compliance

We evaluated exercise adherence based on attendance and intensity compliance.

*Attendance* was defined as the proportion of sessions attended by each patient compared to the total number of sessions offered. The 12-week intervention was planned with three sessions scheduled per week, resulting in a total of 36 sessions, and potential variations due to holidays and logistical issues were taken into account when determining individual attendance rates. Based on thresholds commonly used in cardiac rehabilitation literature, attendance was categorized into three levels: ≥70% (minimum attendance), ≥80% (high attendance), and ≥90% (very high attendance). This categorization allowed for the exploration of potential differences in attendance patterns between intervention groups.

Several strategies were planned to be implemented to boost patient attendance and sustain long-term physical activity levels: (i) Study participants were initially given the choice to select their training session times, with the option to make adjustments as needed during the study. (ii) Automated reminder messages were sent prior to each session, with additional, tailored phone calls made to patients exhibiting inconsistent attendance patterns to foster ongoing engagement. (iii) During the sessions, patients were allowed to listen to music.

Participants were informed at enrolment that, on completing the entire study, they would receive: (i) a personalised report summarising their fitness-assessment results and training data, (ii) a brochure with practical tips for maintaining an active lifestyle, and (iii) a study-branded T-shirt. These items were provided to both the intervention and control groups.

*Intensity compliance* was assessed with two complementary heart-rate–based criteria: first, the second-by-second trace had to show at least thirty seconds over 85 % HRpeak during the first high-intensity interval and at least two minutes in that zone during each subsequent interval, which translates to a minimum accumulated exposure of 6 minutes and 30 seconds per session in the HIIT protocol and 4 minutes and 4 seconds in the HIIT + RT protocol. Second, in accordance with the guidelines developed by Taylor et al. (2019) (32) for each interval we extracted the highest HRpeak rate recorded in its final thirty seconds, averaged these peak values across the four intervals in HIIT or the three intervals in HIIT + RT, and counted the session as compliant when that mean reached or exceeded 85% of HRpeak.

#### Enjoyment and affective response

As part of the intervention assessment, the patients’ enjoyment was evaluated every two weeks using the Physical Activity Enjoyment Scale (PACES) questionnaire (54). The goal of this approach was to evaluate comfort and satisfaction of the patients with the exercise interventions. The scale ranges from 1 (lowest enjoyment) to 7 (highest enjoyment).

The Feeling Scale (43) measured affective responses both prior to and following each exercise session. The Feeling-Scale is a one-dimensional scale consisting of 11 points, spanning from −5 (very poor) to +5 (excellent), with 0 meaning a neutral response. Because the Feeling-Scale is a single-item measure of general mood, the score is not limited to exercise perse, however, by collecting it right before and right after each session we treated the ratings as the participants’ immediate emotional reaction to that particular bout of exercise.

#### Perceived of exertion (RPE)

Immediately after every work bout the supervising exercise trainer asked participants to rate their effort using the Borg category-ratio 0–10 scale. In the HIIT arm, a rating was collected at the end of each of the 4-min intervals. In the HIIT + RT arm, RPE was likewise recorded after the 3 aerobic intervals and after the final repetition of each exercise in the two resistance-training circuits (sets 1 and 2). All RPE values were entered in real time into REDCap by the trainer.

### Statistical Analysis

To test the hypothesis that both interventions (HIIT and HIIT+RT) would result in similar outcomes across all measured variables, we conducted between-group comparisons for session attendance, intensity compliance, perceived exertion, enjoyment, and affective responses.

All analyses were conducted in R version 4.4.0. Data distributions were first inspected using histograms and Q–Q plots and formally assessed for normality with the Shapiro– Wilk test. Between-group comparisons of normally distributed continuous outcomes (e.g., PACES scores, overall attendance percentages, RPE) were carried out using independent two-sample t-tests, applying Welch’s correction when variances were unequal; non-normal continuous variables were compared with the Mann–Whitney U test. Within-subject changes (pre-vs. post-session Feeling Scale scores; RPE between RT sets in the HIIT+RT group) were evaluated by paired t-tests or Wilcoxon signed-rank tests according to the distribution of differences. Proportions of participants meeting attendance thresholds (≥70%, ≥80%, ≥90%) were compared using Pearson’s Chi-squared tests. Missing data were handled using available-case analysis, including all valid observations for each variable or time point. All tests were two-sided and interpreted at a significant level of p < 0.05.

## Results

### Patients’ characteristics

During the 12 weeks 8 participants dropped-out (HIIT group (n=4) and HIIT+RT (n=4), scheduling conflict (n=2) and health problems (n=6)), the final analysis included a total of 64 patients with CAD (HIIT=31 and HIIT+RT=33). The mean age was 62.0 ± 6.7 years, and the mean peak oxygen uptake (VO₂peak) was 25.6 ± 5.0 mL·kg⁻¹·min⁻¹. The percentage of male patients was 77.8%. Based on body mass index (BMI), 9.7% of participants were classified as having normal weight, 55.6% as overweight, and 34.7% as obese. Regarding educational level, 20.8% had completed primary education, 43.1% had completed secondary education, and 36.1% held a university degree. These characteristics are detailed in Table 1.

**Table 1.** Baseline characteristics of participants assigned to the exercise interventions(n = 72)

| Variable | Value |
| --- | --- |
| **n** | 72 |
| Age, years | 62.0 ± 6.7 |
| VO<sub>2</sub>peak, ml·kg<sup>-1</sup>·min<sup>-1</sup> | 25.6 ± 5.0 |
| **Sex, n (%)** |  |
| Male | 56 (77.8) |
| Female | 16 (22.2) |
| **BMI category, n (%)** |  |
| Normal weight | 7 (9.7) |
| Overweight | 40 (55.6) |
| Obesity | 25 (34.7) |
| **Education level, n (%)** |  |
| Primary school | 15 (20.8) |
| Secondary school | 31 (43.1) |
| University | 26 (36.1) |

Values are presented as mean ± standard deviation (SD) or number and percentage (%). VO<sub>2</sub>peak: peak oxygen uptake (in mL·kg<sup>-1</sup>·min<sup>-1</sup>); BMI: body mass index.

### Attendance

Overall, the mean attendance across the sample (n = 64) was 88%, with participants attending an average of 30.0 ± 5.2 sessions out of the 36 scheduled sessions. A total of 60 participants (94%) attended ≥70% of the offered sessions, 53 (83%) attended over 80%, and 33 (52%) attended over 90%, (Figure 3A).

**Fig. 3.**
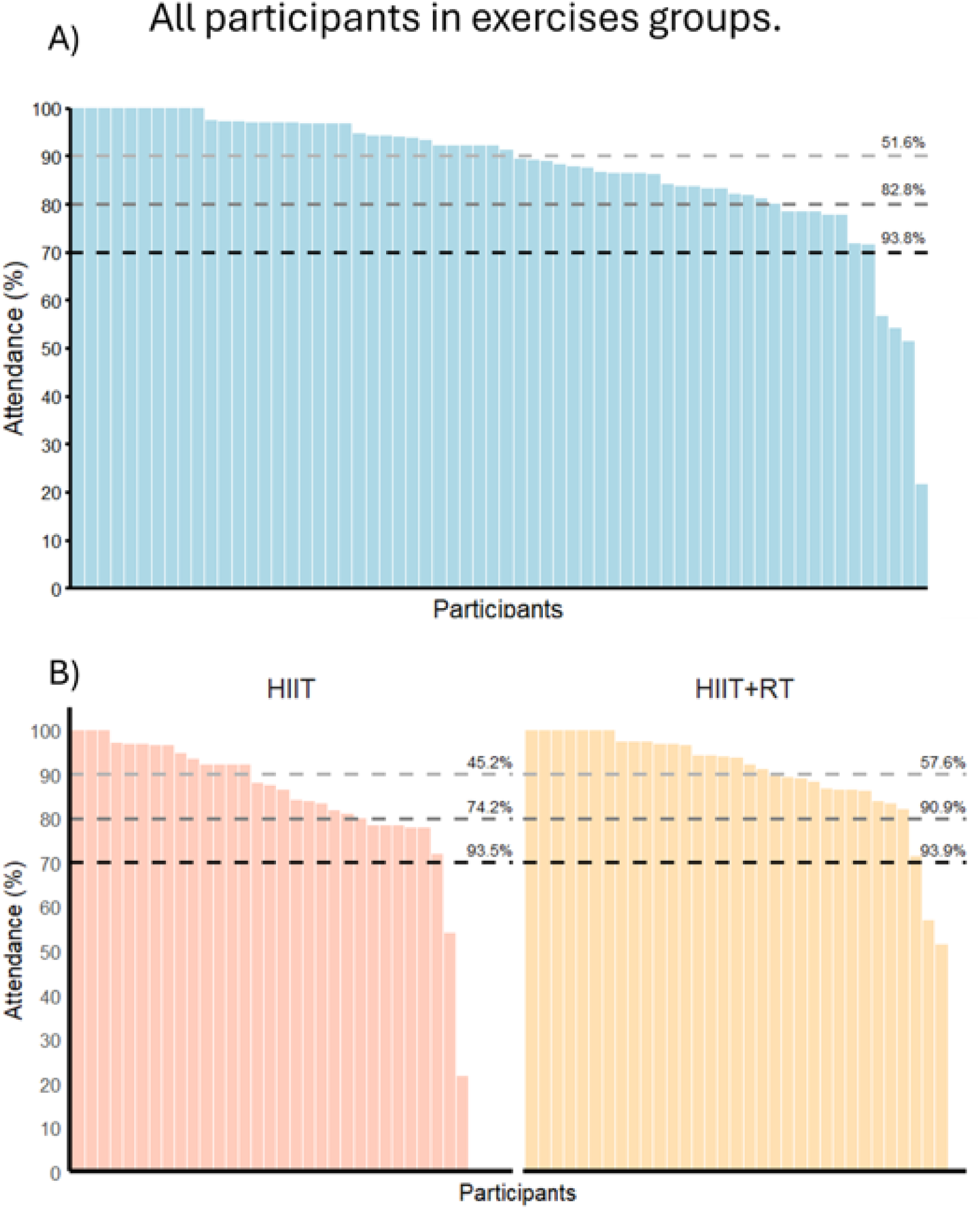
Attendance Rates Across Patients: Total Sample and Stratified by Intervention Groups (HIIT and HIIT+RT). Percentage of patients above attendance thresholds is nested. For example, the percentage of patients making the 70% threshold (i.e. 93.5% of patients in the HIIT group), means they attended 70% of the session or more. The 80% threshold indicates how many patients attended 80% of the sessions or more. The 90% threshold indicates how many patients attended 90% of the sessions or more.

When analyzed by intervention group, the HIIT group had a mean attendance of 29 ± 5 sessions (85%), while the HIIT+RT group attended an average of 31 ± 5 sessions (90%).

The between-group difference in mean attendance was 5% (95% CI [–11.93, 1.79]; p = 0.145).The proportion of participants attending at least 70% of the sessions was nearly identical in both groups, with 94% (29/31) in the HIIT group and 94% (31/33) in the HIIT+RT group. For the 80% attendance threshold, 74% (23/31) of HIIT participants reached this level compared to 91% (30/33) in the HIIT+RT group. At the highest threshold of 90% attendance, 45% (14/31) of HIIT participants and 58% (19/33) of HIIT+RT participants met the criterion. No statistically significant differences were observed between groups across the three attendance thresholds (p = 1.00, p = 0.15, and p = 0.46, respectively).

### Intensity compliance

We analysed second-by-second heart rate data from all available training sessions, resulting in over 3.5 million heart rate records across 64 participants. After excluding non-attended, non-offered, and invalid-HR data sessions, a total of 1,228 valid exercise sessions were included in the intensity compliance analysis. The distribution of training intensity zones (%HRpeak) across both intervention groups can be visualized in Figure 4. Overall, the average intensity compliance rate for the full sample was 75 ± 30%, reflecting the proportion of sessions in which participants met the target heart rate zone criteria.

**Fig. 4.**
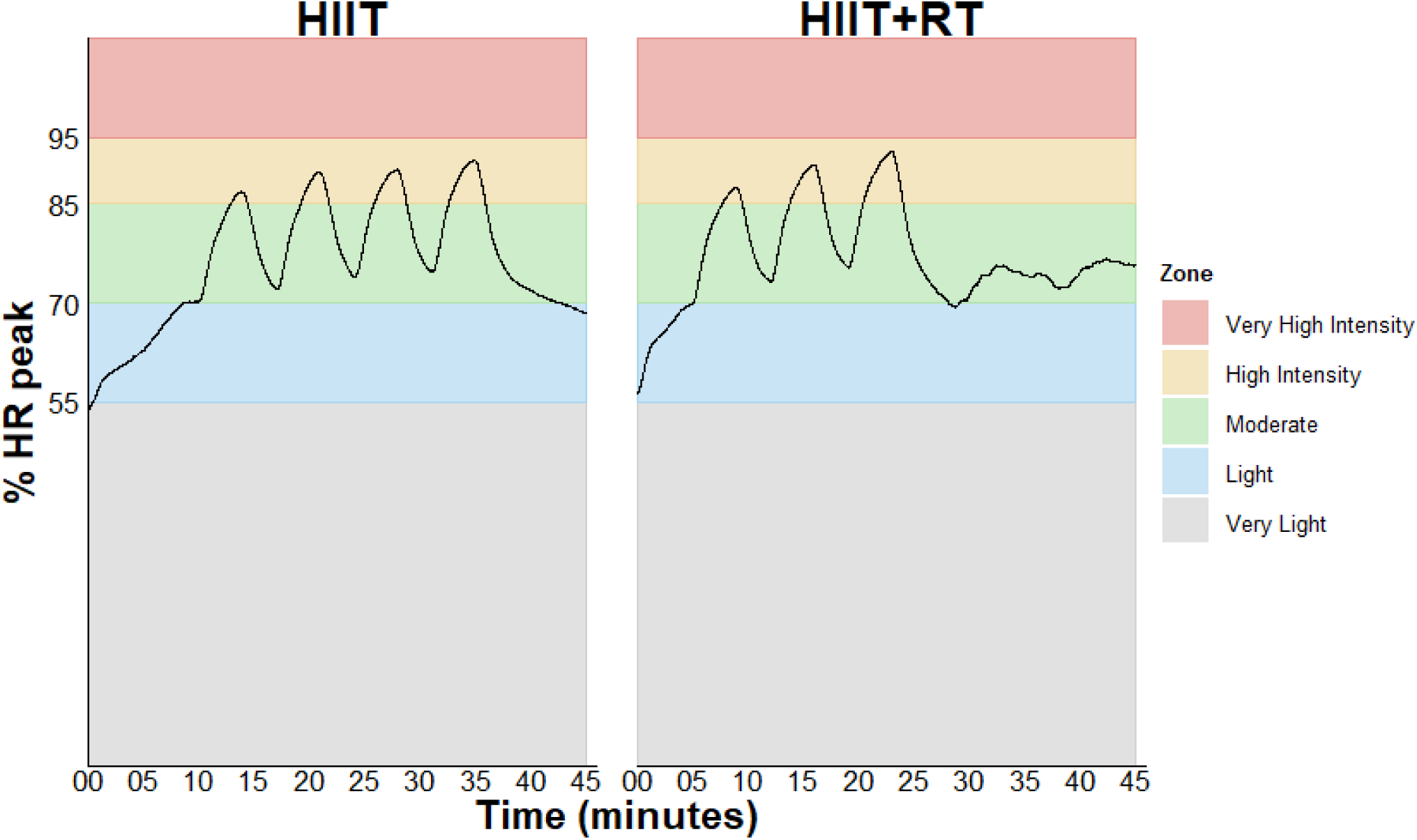
Mean second-by-second heart rate across all sessions (+3.5 million data points analyzed) by intervention group (Group 1: HIIT, Group 2: HIIT+RT), showing time spent in different intensity zones (%HRpeak).

When broken down by intervention group, the HIIT group achieved a mean compliant sessions of 72 ± 33%, while the HIIT+RT group (only the HIIT part of the session analyzed here) showed a slightly higher mean of 78 ± 26%. No statistically significant difference in compliance was observed between intervention groups, with a mean difference of 5% (95% CI: –10 to 20; p = .476), as shown in Figure 5.

**Fig. 5.**
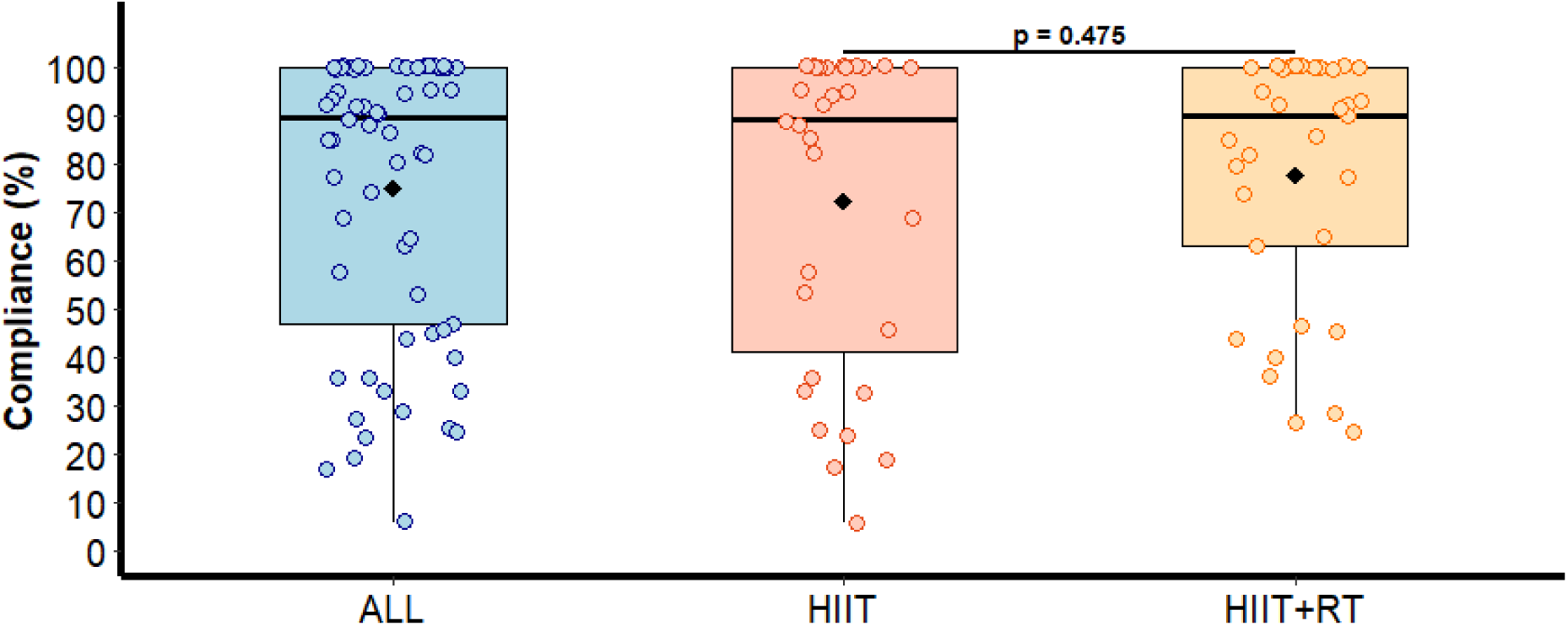
Intensity Compliance Rates Based on Heart Rate Monitoring: Total Sample and Stratified by Intervention Groups (HIIT and HIIT+RT). Box-and-whisker plots show the percentage of completed sessions in which each participant reached ≥ 85 % HRpeak, calculated from second-by-second Polar heart-rate recordings across the whole program. For the HIIT + RT arm, intensity compliance is calculated only for the three aerobic intervals performed within each combined session. black horizontal line = group mean; diamond (rhombus) = median. The p-value (Welch t-test) compares median intensity compliance between the two intervention groups.

A secondary, pragmatic method to assess intensity compliance was also applied, based on the average HR recorded during the high-intensity intervals of each session, as recommended by HIIT exercise guidelines in clincal populations (32). The average intensity compliance across the entire sample was 78 ± 25%, as shown in Supplementary Material Figure 1.

When stratified by intervention group, the HIIT group achieved a mean compliant sessions of 72 ± 30% (n = 31), while the HIIT+RT group reached 84 ± 20% (n = 33). Although intensity compliance tended to be higher in the HIIT+RT group, no statistically significant difference was observed between groups (Welch’s t-test, *p* = 0.13).

### Physical Activity Enjoyment Scale (PACES)

The overall mean PACES score across all patients was 5.4 ± 0.9 points, indicating generally high enjoyment levels during the intervention (Figure 6A). The HIIT group had a mean score of 5.3 ± 0.9, while the HIIT+RT group scored 5.5 ± 0.9. Therefore, a parametric independent samples t-test was applied, revealing no statistically significant difference between groups (p = 0.27).

**Fig. 6.**
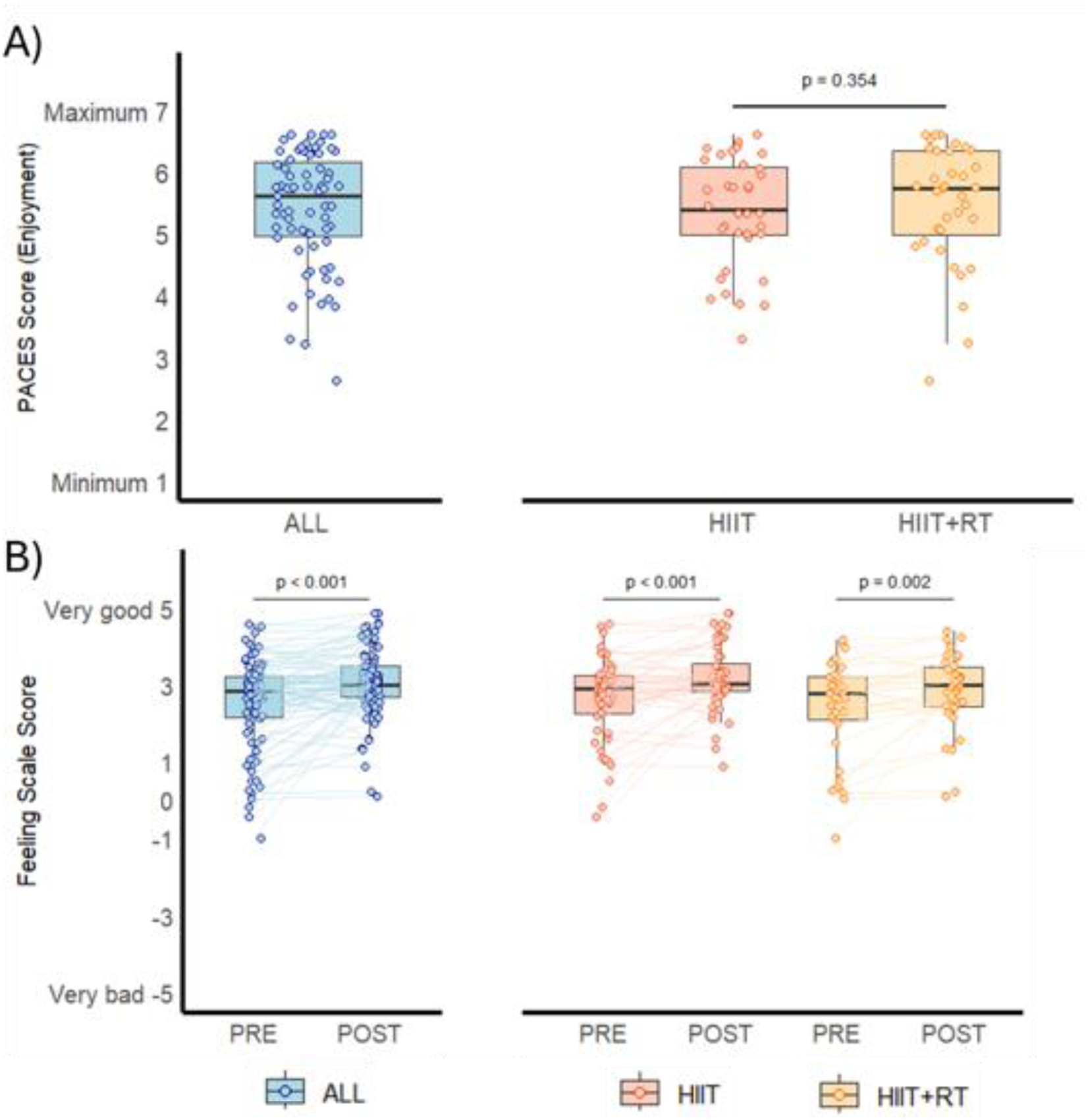
Enjoyment and Affective Responses to Exercise: PACES and Feeling Scale Scores in Total Sample and by Intervention Groups (HIIT and HIIT+RT)

### Feeling Scale

Significant improvements in Feeling-Scale scores were observed post-exercise compared to pre-exercise across all sessions for both groups. For the full sample, the mean Feeling Scale score increased significantly from pre-to post-session (p < 0.001). In the HIIT group, scores improved from 2.6 ± 1.2 pre-session to 3.3 ± 0.9 post-session (p < 0.001), while in the HIIT+RT group, scores increased from 2.4 ± 1.3 to 2.9 ± 1.00 (p = 0.002) (Figure 6B). An ANCOVA was conducted to compare post-exercise Feeling Scale scores between intervention groups while controlling baseline (pre-exercise) values. After adjusting for pre-session scores, there was a statistically significant effect of the intervention group on post-session scores, (F(1, 2373) = 92.6, p < .001), indicating that participants in the HIIT+RT group reported significantly more positive affective responses following exercise compared to those in the HIIT group.

### Perceivded Exertion (RPE)

The distribution of RPE scores was examined for normality using the Shapiro–Wilk test and graphical methods. During the high-intensity intervals, the overall mean RPE was 7.1 ± 0.7, reflecting moderate-to-high exertion levels. The HIIT group reported a mean of 7.1 ± 0.7, and the HIIT+RT group a mean of 7.1 ± 0.6. The between-group difference was negligible and not statistically significant (mean difference = 0.0, 95% CI [–0.2, 0.2], *p* = 0.987; independent *t*-test; Supplementary Material Figure 2A). In the resistance training part of the HIIT+RT sessions, the mean RPE increased from 5.0 ± 0.9 in Set 1 to 5.2 ± 0.8 in Set 2. This difference was statistically significant (mean difference = 0.2, 95% CI [–0.1, 0.4], *p* = 0.02; paired *t*-test; Supplementary Material Figure 2B).

## Discussion

The aims of this study were to (i) provide a CERT-based description of the two HIIT-based exercise interventions implemented in the Heart-Brain study and (ii) quantify and compare the adherence, intensity compliance, enjoyment and affect response to the programs. Our key findings were that there were no differences between HIIT and HIIT+RT in terms of adherence (attendance of sessions), intensity compliance with the prescribed heart-rate zones, perceived effort during the high-intensity bouts, or enjoyment. Both intervention groups also showed affective tone improved within every session as reflected by Feeling-Scale scores significantly improving from pre to post session.

Both of the Heart-Brain exercise programs demonstrated high adherence with participants attending 88% of all scheduled sessions, and complied with the prescribed intensity 75% of the time. These adherence rates are broadly consistent with previous studies, such as Kristiansen *et al.*’s HIIT trial (91 % attendance; 86 % compliance) (22), the FITR-Heart RCT (83 % attendance; 72 % compliance) (55), and the HIITERGY study (≈ 90 % attendance; 73 % compliance) (56). These results are higher than the 64% attendance reported in a large U.K. phase-III CR programme (17). Because exercising in the target zone is a pre-requisite for optimal cardiorespiratory adaptation (57), guidelines to combine objective HR control with the practical guidance of RPE (32) appears to be a pragmatic strategy for routine care.

Enjoyment was consistently high with a mean PACES score of 5.3 for HIIT and 5.5 for HIIT + RT on a 1-to-7 scale. This mirrors the ∼5.5/7 reported by Gayda et al. in CAD patients performing interval sessions (23) and exceeds the ≥4.5 threshold associated with future exercise uptake (12). Feeling-Scale scores also improved during the sessions (before vs. after). In the HIIT arm the mean score increased by +0.7 points, while the HIIT + RT arm increased by +0.5 points. These changes are comparable to the +0.5-point improvement reported in Kristiansen’s HIIT trial (22) and the +0.4-point increase observed by Bartlett et al. during interval running (58). Such affective responses have been associated with better long-term adherence (13,14,57,59,60).

Perceived exertion during the 4-minute high-intensity intervals was consisntenly within the target “hard” zone for both interventions with a mean RPE of 7.1. This aligns with the RPE of 7.0 reported by Kristiansen et al. in HIIT-trial (22) and the 16.3 (using the 6-20 RPE) seen in the FITR-Heart trial (55), confirming that our protocol met the clinical recommendation to reach an RPE of seven or higher (15,32).

Within the resistance-training portion of the HIIT + RT sessions, perceived effort began at 5.0 on the Borg 10 scale during the first circuit and increased to 5.2 in the second. An exertion of five corresponds to “somewhat hard” work and sits squarely in the window recommended by the American College of Sports Medicine and recent clinical reviews (approximately five to six) for delivering at least sixty per cent of one-repetition-maximum without undue cardiovascular strain (15,19). Notably, these values replicate the 4.9 reported for strength circuits embedded in whole-body HIIT for coronary patients by Kristiansen et al. (22) and the pooled RPE of 5 summarised across eleven CAD resistance-training trials in the meta-analysis by Hollings and colleagues (61). They are consistent with the RPE 5.1 observed in high-effort resistance protocols reviewed by Steele et al. (26). The modest but significant rise between circuits likely reflects normal fatigue-related drift rather than excessive overload, underscoring that two short strength circuits can be layered onto a 4-interval HIIT session without compromising either safety or the intended training intensity in cardiac rehabilitation.

Our research found that individuals in the HIIT+RT intervention demonstrated numerically higher attendance and intensity compliance compared to those in the HIIT-only group, although not statistically significant. Enjoyment was also slightly higher in the HIIT+RT group, with a mean PACES score of 5.5 versus 5.3 in the HIIT group, yet differences were not statistically significant either. This may be partially explained by several factors. First, the inclusion of RT introduced greater variety within the sessions, potentially making the experience more engaging and less monotonous (61). Additionally, participants may have perceived the combination of aerobic and strength training as a more complete or functional workout, increasing their motivation and perceived value of the program (62). The HIIT + RT sessions also involved one fewer high-intensity interval, which may have reduced overall cardiovascular fatigue and thereby improved tolerability (32). For some individuals, the strength component might have been more appealing and/or familiar, especially among those with prior gym experience, enhancing their willingness to attend and complete sessions (61).

From a practical perspective, this research provides empirical evidence supporting the feasibility of integrating HIIT and HIIT+RT in supervised exercise programmes for patients with CAD, while highlighting the need for standardised reporting to ensure reproducibility and facilitate effective implementation in clinical practice. By adhering to the CERT guidelines (30), our study ensures a transparent and standardised description of the interventions, facilitating its replication and broader applicability in real-world rehabilitation programs (63,64). Given the existing variability in the reporting of exercise interventions, particularly in terms of intensity prescriptions and session structure, the adoption of standardised frameworks like CERT is crucial to ensuring consistency across studies and optimizing patient outcomes (63,64). Our findings reinforce the importance of precise intervention reporting, allowing exercise programs to refine their protocols based on structured HIIT and HIIT+RT regimens while ensuring that exercise prescriptions are clearly defined, monitored, replicable, and aligned with best clinical practices.

The study has several limitations that must be acknowledged. While the intervention demonstrated high attendance and intensity compliance over 12 weeks, long-term adherence to exercise beyond the study period was not assessed. Future studies should incorporate longitudinal follow-ups (e.g., 6 months or 1 year post-intervention) to evaluate whether the observed engagement levels and intensity compliance translate into sustained physical activity behaviors. Second, the absence of a MICT group limits direct comparisons with traditional rehabilitation approaches. Although HIIT is gaining clinical acceptance, examining how MICT, HIIT, and HIIT+RT compare in long-term attendance and physiological outcomes remains an important avenue for research. Trials usually have participation bias, with healthier and more motivated participants being included, which can positively influence the results presented compared to the whole CAD population. Finally, the study was conducted in a single-center setting, with a homogeneous sample of stable CAD patients, which may limit the generalizability of findings to more diverse populations with cardiovascular diseases or individuals with a higher cardiovascular risk. Future multicenter trials with more heterogeneous cohorts would help validate these results in broader clinical settings.

Key strenghs of this study include: i) its innovative approach by implementing two distinct exercise interventions aligned with the latest guidelines (32,33,65,66); ii) its multidisciplinary design, in which researchers, cardiologists, exercise trainers, and cardiac nurses worked together to provide a fully documented, structured HIIT and HIIT+RT protocols that can serve as a model for future clinical applications; iii) the comprehensive assessment of attendance, intensity compliance, enjoyment and affective responses; iv) the use of second-by-second HR tracking to assess intensity compliance with intensity prescriptions, analyzing more than 3.5 million data points of seconds; v) the comparison of two exercise modalities (HIIT and HIIT+RT) with equivalent total workload and session duration strengthens the study’s internal validity; and vi) its rigorous design and comprehensive reporting following CERT guidelines, enhancing transparency and replicability, key aspects that are often insufficiently described in exercise trials.

### Conclusions

This study shows that two CERT-described, guideline-aligned protocols (HIIT and HIIT + RT) are both feasible and well-accepted by patients with coronary artery disease. An overall high attendance rate of 88%, heart-rate–verified intensity compliance of 75%, and high enjoyment scores in both arms, confirms that favourable affective responses can coexist with vigorous training stimuli. The HIIT + RT format, which integrates strength work without extending total session time, offers an attractive option for routine clinical programmes and future trials aimed at maximising cardiometabolic and musculoskeletal benefits within limited rehabilitation schedules. By presenting the interventions with full CERT transparency, we also provide a replicable template that can accelerate the translation of exercise science into day-to-day clinical practice.

## Data Availability

All data produced in the present study are available upon reasonable request to the authors

## Ethical Approval

The trial protocol is in accordance with the principles of the Declaration of Helsinki and was approved by the Research Ethics Board of the Andalusian Health Service (CEIM/CEI Provincial de Granada; #1776-N-21on December 21st, 2021).

## Supplementary material

### Supplementary Material 1.

**Table 1.** CERT checklist from the Heart-Brain study exercise intervention.

|  | Item | Checklist item | Identification, section |
| --- | --- | --- | --- |
| What:<br>Materials | 1 | Detailed description of the type of exercise equipment | Methods - Location, providers, and materials (Second paragraph), Supplementary Material Table 4 |
| Who:<br>Provider | 2 | Detailed description of the qualifications, expertise and/or training | Methods - Location, providers, and materials (First paragraph mentions recording of attendance and monitoring strategies) |
| How:<br>Delivery | 3 | Describe whether exercises are performed individually or in a group | Methods - Delivery of the training (First and second paragraph) |
|  | 4 | Describe whether exercises are supervised or unsupervised; how they are delivered | Methods - Delivery of the training (First paragraph) |
|  | 5 | Detailed description of how adherence to exercise is measured and reported | Methods - Adherence, attendance and intensity compliance (First paragraph, mentions recording of attendance and monitoring strategies) |
|  | 6 | Detailed description of motivation strategies | Methods - Adherence, attendance and intensity compliance (Second paragraph) |
|  | 7a | Detailed description of the decision rule(s) for determining exercise progression | Methods - Delivery of the training (Fourth paragraph) |
|  | 7b | Detailed description of how the exercise program was progressed | Methods - Delivery of the training (Fourth paragraph) |
|  | 8 | Detailed description of each exercise to enable replication | Methods - HIIT-based intervention structure, Supplementary Material Table 2 (Detailed exercise structure) & Supplementary Material Table 3 for the resistance-training circuit. |
|  | 9 | Detailed description of any home program component | Home-based exercise intervention was not included in the intervention design |
|  | 10 | Describe whether there are any non-exercise components | No additional non-exercise components were included in the intervention, as the focus was exclusively on supervised exercise sessions. |
|  | 11 | Describe the type and number of adverse events that occur during exercise | Methods - Safety and adverse events |
| Where:<br>location | 12 | Describe the setting in which the exercises are performed | Methods - Location, providers, and materials (First paragraph) |
| When:<br>how much:<br>dosage | 13 | Detailed description of the exercise intervention | Methods - Exercise sessions characteristics (First paragraph, detailed session structure and duration); Methods - Description of HIIT-based exercise intervention - Exercise dosage and isoenergetic calculation (exercise frequency, intensity, and energy expenditure); Supplementary Material Tables 2 5 and 6 |
| Tailoring:<br>what, how | 14a | Describe whether the exercises are generic (one size fits all) or tailored | Methods - Delivery of the training (Second paragraph.) |
|  | 14b | Detailed description of how exercises are tailored to the individual | Methods - Delivery of the training (Second paragraph) |
|  | 15 | Describe the decision rule for determining the starting level | Methods - Delivery of the training (Second paragraph) |
| How well:<br>planned,<br>actual | 16a | Describe how adherence or fidelity is<br>assessed/measured | Methods - Adherence (Allparagraphs) |
|  | 16b | Describe the extent to which the intervention was<br>delivered as planned | Results – Intensity Compliance (First<br>paragraph); Methods – Intensity<br>Compliance assessment; Supplementary<br>Material Figure 1. |

**Table 2.** Description detailed of the exercises

*This table includes photographs of individuals whose inclusion is incompatible with the rapid nature of preprint posting. Please contact the corresponding author to request access to these images*.

**Table 3.** Exercises and patterns of movement of the resistance part.

| *Number of*<br>*exercises* | *Pattern of*<br>*movement* | *Exercise* |
| --- | --- | --- |
| 1<sup>o</sup> | Vertical traction | Pull over or<br>Pull down |
| 2<sup>o</sup> | Core | Front plank or<br>Lateral plank |
| 3<sup>o</sup> | Vertical push | Shoulder press |
| 4<sup>o</sup> | Legs | Squat |
| 5<sup>o</sup> | Horizontal traction | Row |
| 6<sup>o</sup> | Horizontal push | Push-ups |
| 7<sup>o</sup> | Core | Press pallof |
| 8<sup>o</sup> | Legs | Bridge or<br>Lunge |

**Table 4.**
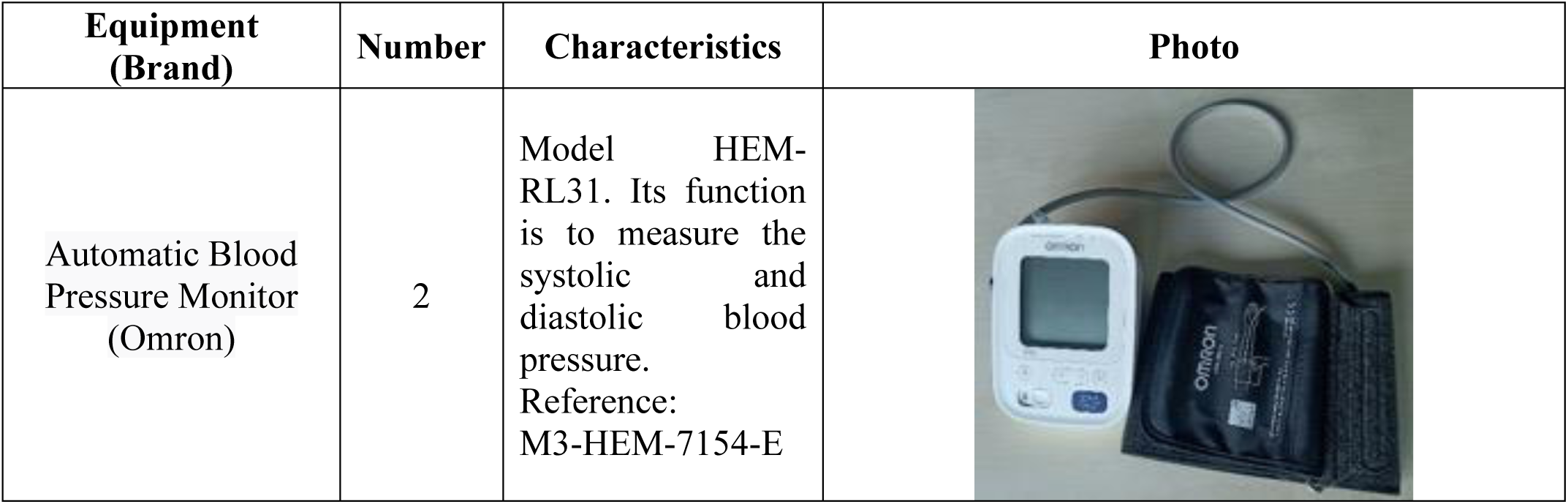
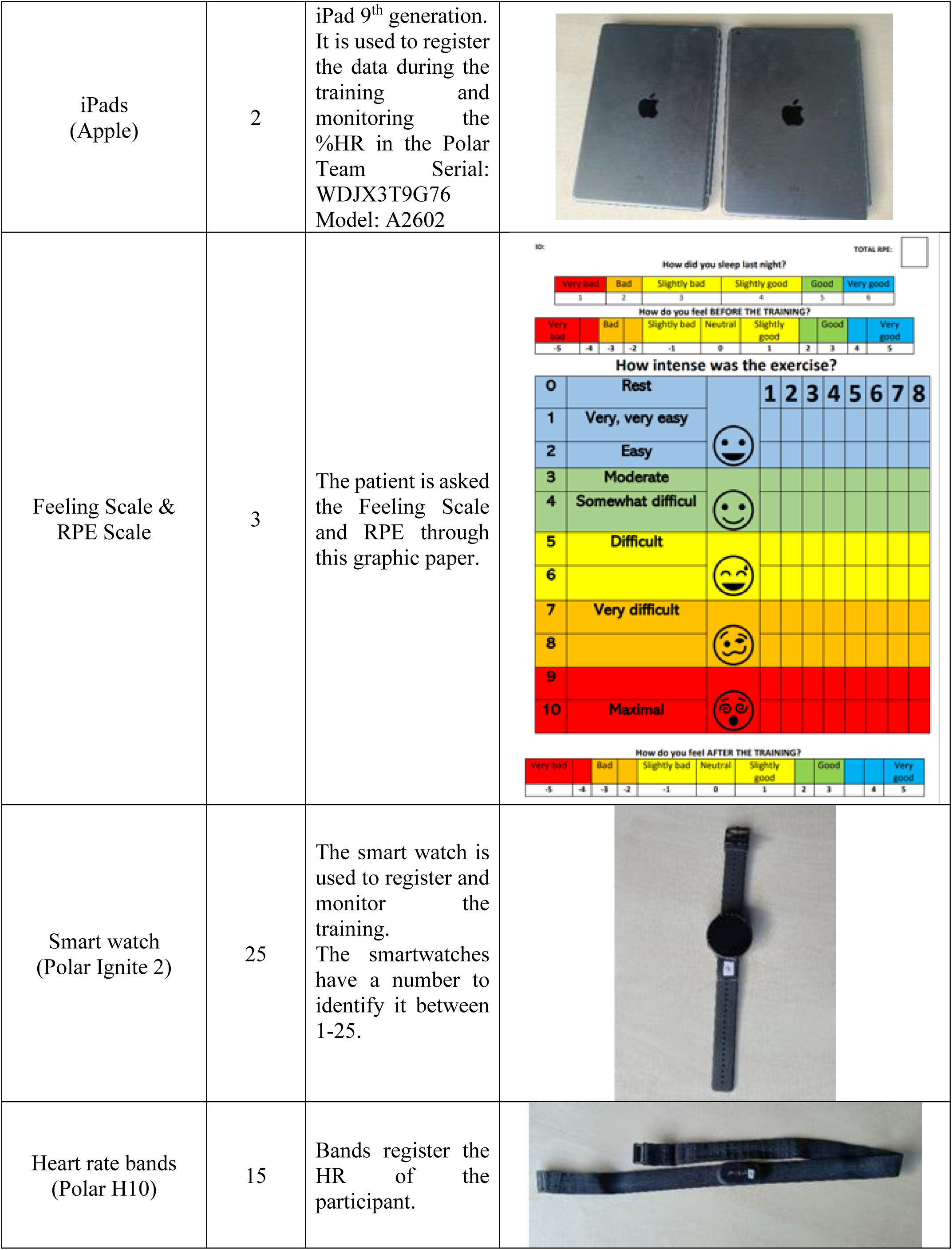
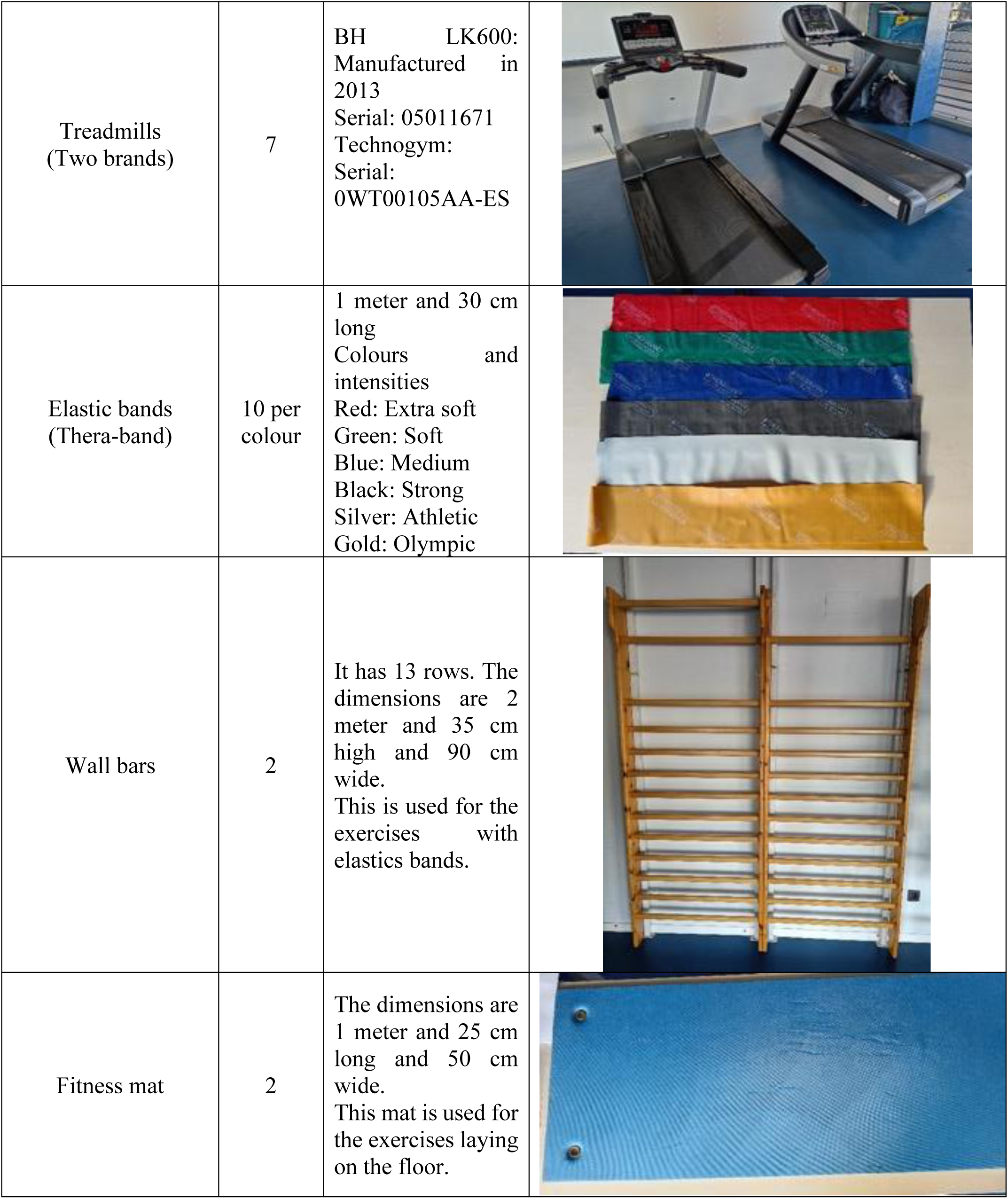
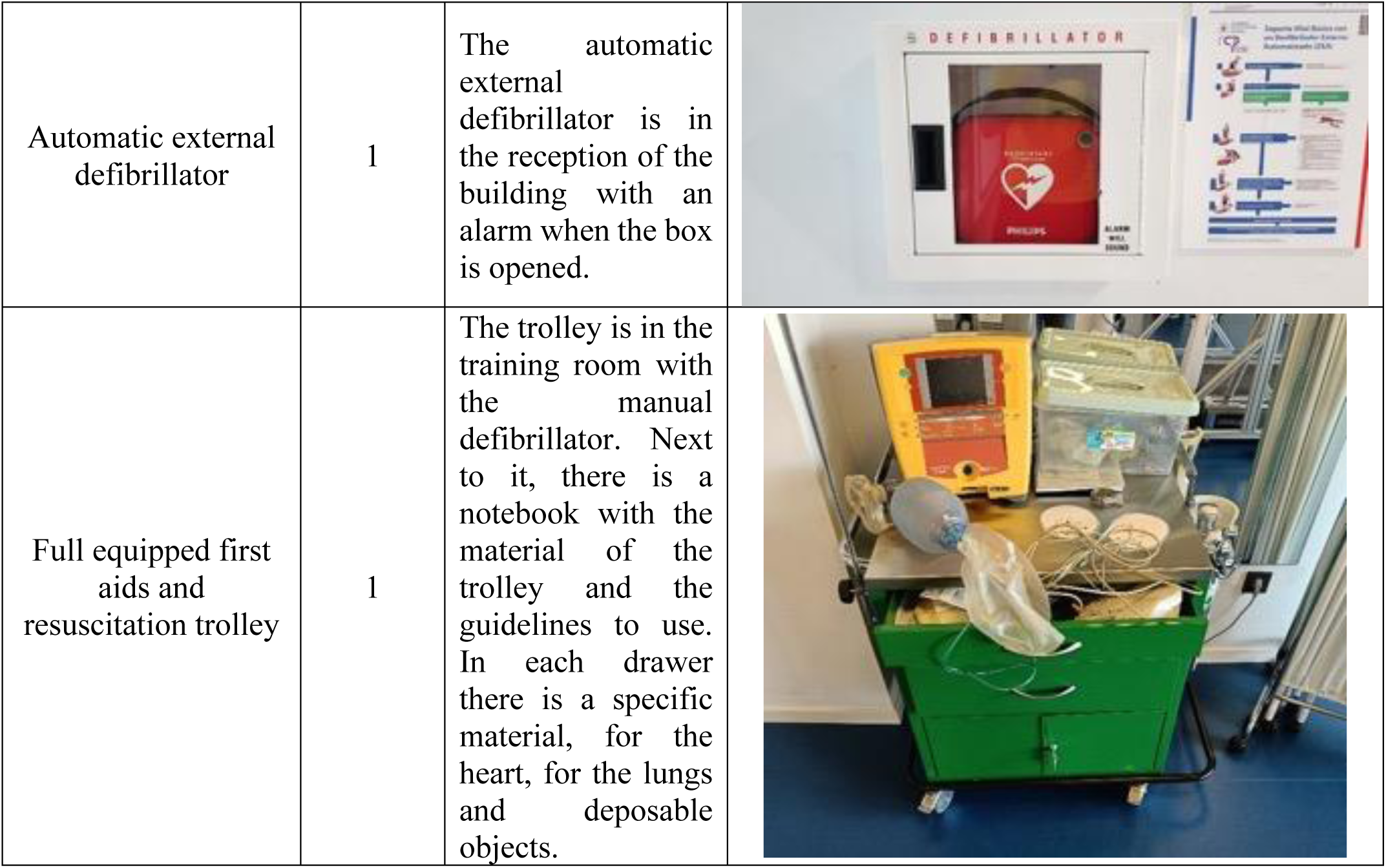
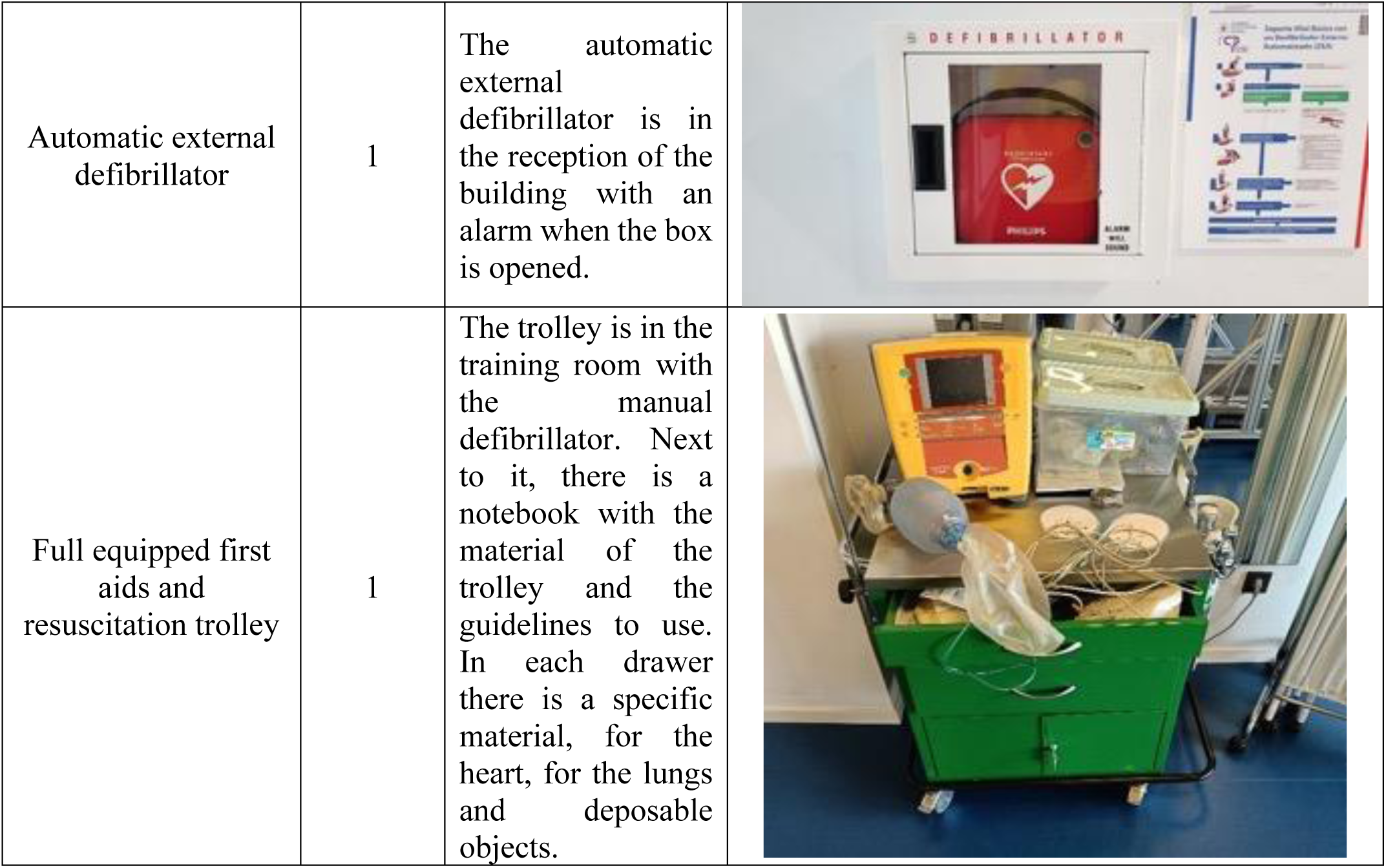
Exercise equipment.

**Table 5.** Relation of zone of intensity and %HR peak.

| *Number* | *Intensity* | *%HR peak* |
| --- | --- | --- |
| 1 | Very light | 1-50 |
| 2 | Light | 51-65 |
| 3 | Moderate | 66-84 |
| 4 | High-intensity | 85-95 |
| 5 | Very high-intensity | 96-100 |

**Table 6.** TRIMP calculations (adapted) method, (TRIMP= training impulse)

| **HIIT** | **Time**<br>**(minutes)** | **Intensity zone** | **TRIMP**<br>**(Time \* Intensity zone)** | **TRIMP/week** | **TRIMP per**<br>**minute**<br>**(TRIMP/session**<br>**time)** |
| --- | --- | --- | --- | --- | --- |
| **Warm-up** | 10 | 3 | 30 | 90 | 3 |
| **High intensity**<br>**intervals** | 16 | 4 | 64 | 192 | 4 |
| **Resting**<br>**intervals** | 9 | 3 | 27 | 81 | 3 |
| **Cool-down** | 10 | 3 | 30 | 90 | 3 |
| **Total** | 45 |  | 151 | 453 | 3,35 |
| **HIIT + RT** |  |  |  |  |  |
| **Warm-up** | 5 | 3 | 15 | 45 | 3 |
| **High intensity intervals** | 12 | 4 | 48 | 144 | 4 |
| **Resting intervals** | 6 | 3 | 18 | 54 | 3 |
| **Cool-down** | 5 | 3 | 15 | 45 | 3 |
| **Resistance exercises** | 17 | 3 | 51 | 153 | 3 |
| **Total** | 45 |  | 147 | 452,97 | 3,25 |

**Table 7.** METs calculation per phase of the training, per session and per week of both intervention groups, matching in time and energy expenditure.

| **HIIT** | **Time**<br>**(minutes)** | **METs**<br>**(per intensity)** | **METs / session** | **METs / week** |
| --- | --- | --- | --- | --- |
| Warm-up | 10 | 4.5 | 45 | 135 |
| High-intensity intervals | 16 | 8 | 128 | 384 |
| Resting intervals | 9 | 4.5 | 40.5 | 121.5 |
| Cool-down | 10 | 4.5 | 45 | 135 |
| Total | 45 |  | 258.5 | 775.5 |
| **HIIT + RT** |  |  |  |  |
| Warm-up | 5 | 4.5 | 22.5 | 67.5 |
| High intensity intervals | 12 | 8 | 96 | 288 |
| Resting intervals | 6 | 4.5 | 27 | 81 |
| Cool-down | 5 | 4.5 | 22.5 | 67.5 |
| Resistance exercises | 17 | 5.3 | 90.1 | 270.3 |
| Total | 45 |  | 258.1 | 774.3 |

**Figure s1.**
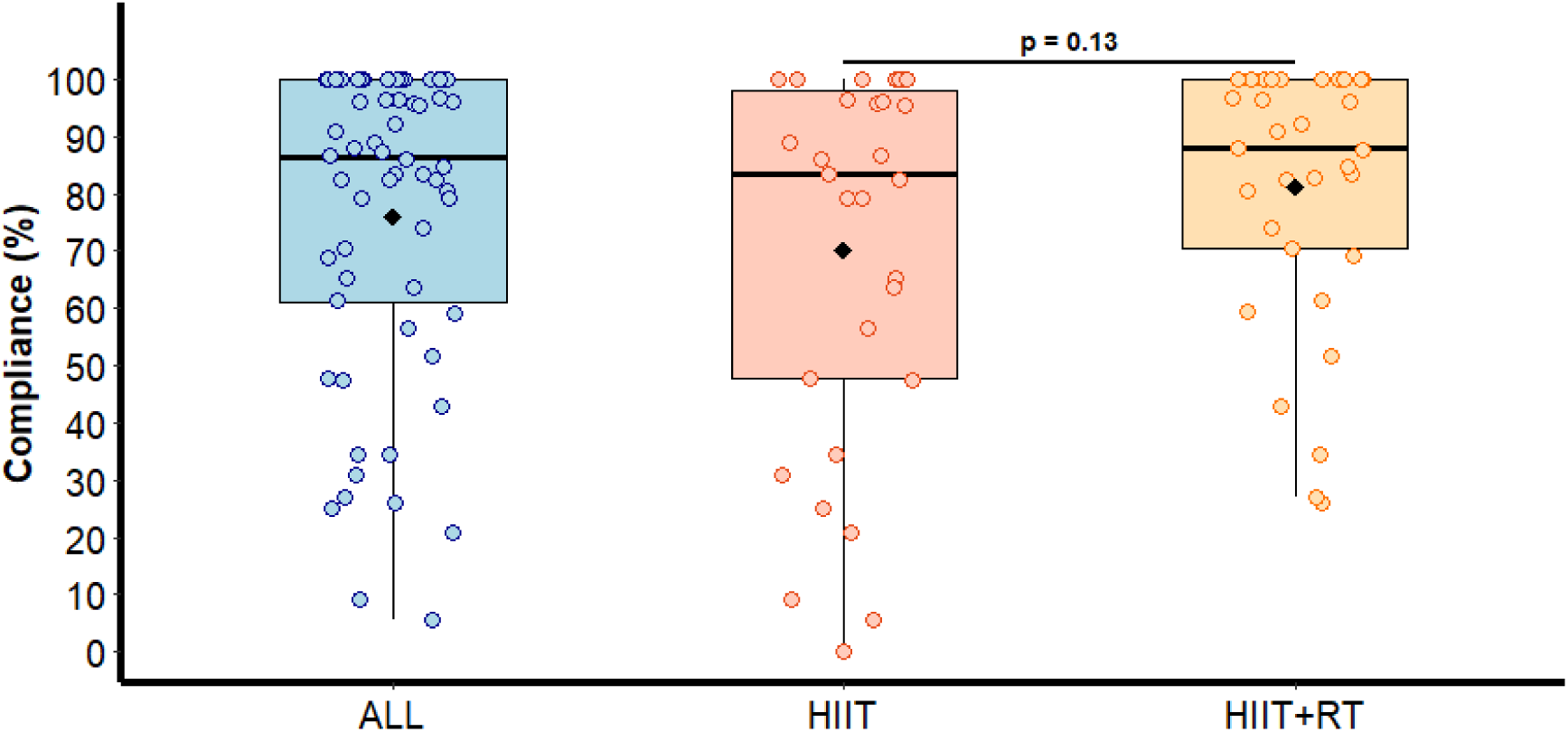
Intensity Compliance Rates Based on Heart Rate Monitoring Using Secondary method (The secondary method evaluated session intensity by capturing%HRpeak during the final 30 seconds of each high-intensity interval). The average of these recorded values was subsequently computed for each session. The session was deemed compliant if the average peak heart rate exceeded 85%, and non-compliant otherwise): Total Sample and Stratified by Intervention Groups (HIIT and HIIT+RT).

**Figure s2.**
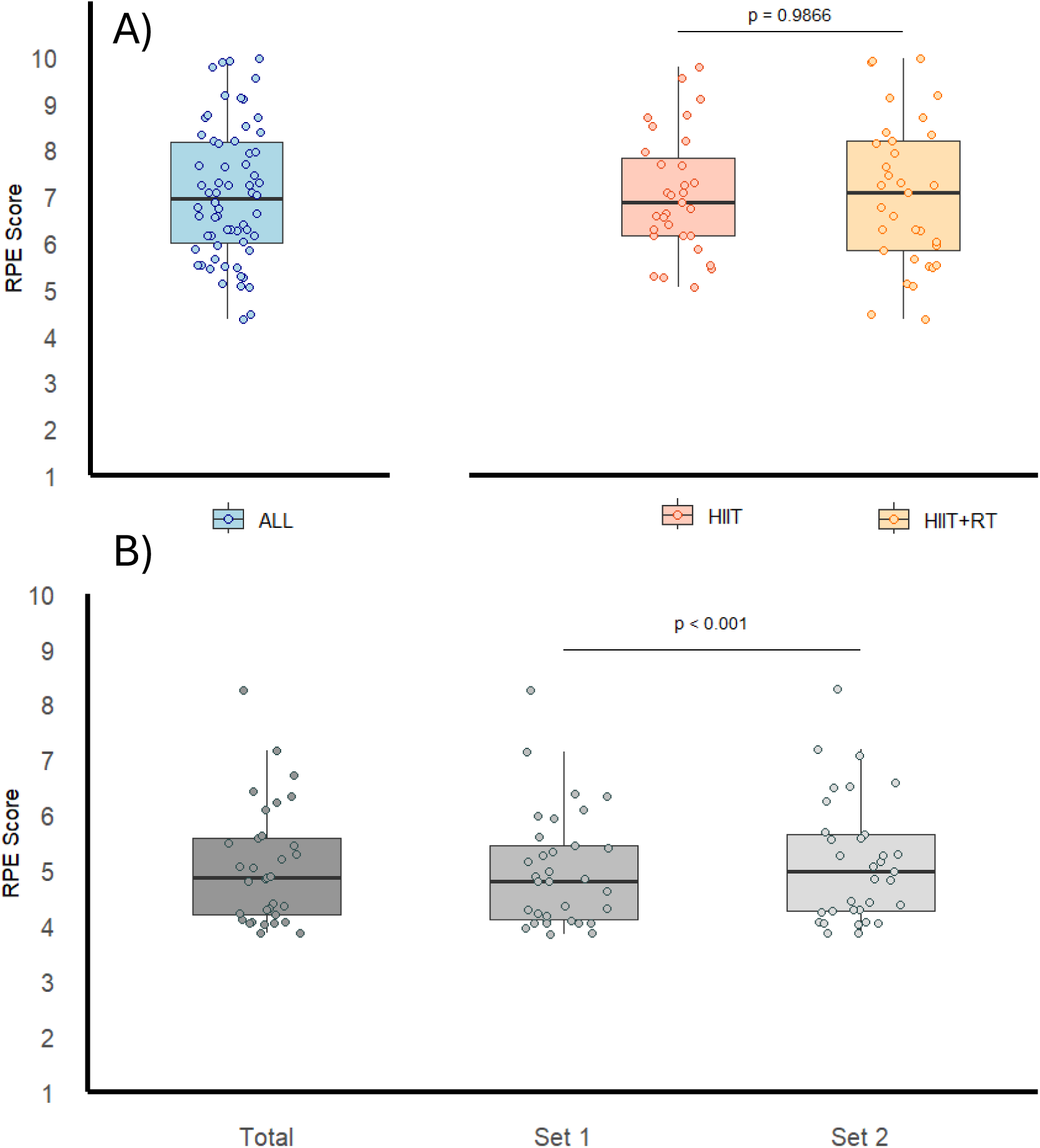
Rate of Perceived Exertion (RPE) During Exercise Sessions: A) High-Intensity Intervals and B) Resistance Training Components.

## References

1. Abbafati C, Abbas KM, Abbasi-Kangevari M, Abd-Allah F, Abdelalim A, Abdollahi M, et al. Global burden of 369 diseases and injuries in 204 countries and territories, 1990–2019: a systematic analysis for the Global Burden of Disease Study 2019. Lancet. 2020;396(10258):1204–22.

2. Gheorghe A, Griffiths U, Murphy A, Legido-Quigley H, Lamptey P, Perel P. The economic burden of cardiovascular disease and hypertension in low- and middle-income countries: A systematic review. BMC Public Health. 2018;18(1):1–11.

3. Virani SS, Newby LK, Arnold S V., Bittner V, Brewer LPC, Demeter SH, et al. 2023 AHA/ACC/ACCP/ASPC/NLA/PCNA Guideline for the Management of Patients With Chronic Coronary Disease: A Report of the American Heart Association/American College of Cardiology Joint Committee on Clinical Practice Guidelines. Vol. 148, Circulation. 2023. 9–119 p.

4. Katta N, Loethen T, Lavie CJ, Alpert MA. Obesity and Coronary Heart Disease: Epidemiology, Pathology, and Coronary Artery Imaging. Curr Probl Cardiol. 2021;46(3):11–20.

5. Taylor RS, Dalal HM, McDonagh STJ. The role of cardiac rehabilitation in improving cardiovascular outcomes. Nat Rev Cardiol. 2022;19(3):180–94.

6. Toval A, Bakker EA, Granada-Maia JB, Núñez de Arenas-Arroyo S, Solis-Urra P, Eijsvogels TMH, et al. Exercise type and settings, quality of life, and mental health in coronary artery disease: a network meta-analysis. Eur Heart J [Internet]. 2025;1–16. Available from: http://www.ncbi.nlm.nih.gov/pubmed/39809303

7. Vonk T, Maessen MFH, Hopman MTE, Snoek JA, Aengevaeren VL, Franklin BA, et al. Temporal trends in cardiac rehabilitation participation and its core components: A nationwide cohort study from the netherlands. J Cardiopulm Rehabil Prev. 2024;44(3):180–6.

8. Ritchey MD, Maresh S, Mcneely J, Shaffer T, Jackson SL, Keteyian SJ, et al. Tracking Cardiac Rehabilitation Participation and Completion Among Medicare Beneficiaries to Inform the Efforts of a National Initiative. Circ Cardiovasc Qual Outcomes. 2020;13(1):E005902.

9. Ruano-Ravina A, Pena-Gil C, Abu-Assi E, Raposeiras S, van ’t Hof A, Meindersma E, et al. Participation and adherence to cardiac rehabilitation programs. A systematic review. Int J Cardiol [Internet]. 2016;223:436–43. Available from: 10.1016/j.ijcard.2016.08.120

10. Resurrección DM, Moreno-Peral P, Gómez-Herranz M, Rubio-Valera M, Pastor L, Caldas de Almeida JM, et al. Factors associated with non-participation in and dropout from cardiac rehabilitation programmes: a systematic review of prospective cohort studies. Eur J Cardiovasc Nurs. 2019;18(1):38–47.

11. Bakhshayeh S, Sarbaz M, Kimiafar K, Vakilian F, Eslami S. Barriers to participation in center-based cardiac rehabilitation programs and patients’ attitude toward home-based cardiac rehabilitation programs. Physiother Theory Pract [Internet]. 2021;37(1):158–68. Available from: 10.1080/09593985.2019.1620388

12. Santaularia N, Jaarsma T. Motivational factors for exercise in cardiac patients? A literature review. Eur J Prev Med. 2013;1(1):20.

13. Lewis BA, Williams DM, Frayeh A, Marcus BH. Self-efficacy versus perceived enjoyment as predictors of physical activity behaviour. Psychol Heal. 2016;31(4):456–69.

14. Klompstra L, Deka P, Almenar L, Pathak D, Muñoz-Gómez E, López-Vilella R, et al. Physical activity enjoyment, exercise motivation, and physical activity in patients with heart failure: A mediation analysis. Clin Rehabil. 2022;36(10):1324– 31.

15. Sabbahi A, Canada JM, Babu AS, Severin R, Arena R, Ozemek C. Exercise training in cardiac rehabilitation: Setting the right intensity for optimal benefit. Prog Cardiovasc Dis [Internet]. 2022;70:58–65. Available from: 10.1016/j.pcad.2022.02.001

16. Taylor JL, Bonikowske AR, Olson TP. Optimizing Outcomes in Cardiac Rehabilitation: The Importance of Exercise Intensity. Front Cardiovasc Med. 2021;8(September):1–17.

17. Khushhal A, Nichols S, Carroll S, Abt G, Ingle L. Insufficient exercise intensity for clinical benefit? Monitoring and quantification of a community-based Phase III cardiac rehabilitation programme: A United Kingdom perspective. PLoS One. 2019;14(6):1–12.

18. De Fazio R, Mastronardi VM, De Vittorio M, Visconti P. Wearable Sensors and Smart Devices to Monitor Rehabilitation Parameters and Sports Performance: An Overview. Sensors. 2023;23(4).

19. of Sports Medicine AC, Liguori G, Fountaine CJ, Feito Y, Roy B. ACSM’s Guidelines for Exercise Testing and Prescription [Internet]. Wolters Kluwer; 2021. (American College of Sports Medicine Series). Available from: https://books.google.es/books?id=hTKvzgEACAAJ

20. Sebastian LA, Reeder S, Williams M. Determining target heart rate for exercising in a cardiac rehabilitation program: A retrospective study. J Cardiovasc Nurs. 2015;30(2):164–71.

21. Hansford HJ, Wewege MA, Cashin AG, Hagstrom A, Clifford BK, McAuley J, et al. If exercise is medicine, why don’t we know the dose? An overview of systematic reviews assessing reporting quality of exercise interventions in health and disease. Br J Sports Med. 2022;692–700.

22. Kristiansen J, Sjúrðarson T, Grove EL, Rasmussen J, Kristensen SD, Hvas AM, et al. Feasibility and impact of whole-body high-intensity interval training in patients with stable coronary artery disease: a randomised controlled trial. Sci Rep [Internet]. 2022;12(1):1–12. Available from: 10.1038/s41598-022-21655-w

23. Gayda M, Ribeiro PAB, Juneau M, Nigam A. Comparison of Different Forms of Exercise Training in Patients With Cardiac Disease: Where Does High-Intensity Interval Training Fit? Can J Cardiol [Internet]. 2016;32(4):485–94. Available from: 10.1016/j.cjca.2016.01.017

24. Wen D, Utesch T, Wu J, Robertson S, Liu J, Hu G, et al. Effects of different protocols of high intensity interval training for VO2max improvements in adults: A meta-analysis of randomised controlled trials. J Sci Med Sport [Internet]. 2019;22(8):941–7. Available from: 10.1016/j.jsams.2019.01.013

25. Ballesta García I, Rubio Arias JÁ, Ramos Campo DJ, Martínez González-Moro I, Carrasco Poyatos M. High-intensity Interval Training Dosage for Heart Failure and Coronary Artery Disease Cardiac Rehabilitation. A Systematic Review and Meta-analysis. Rev Esp Cardiol. 2019;72(3):233–43.

26. Steele J, Plotkin D, Van Every D, Rosa A, Zambrano H, Mendelovits B, et al. Slow and Steady, or Hard and Fast? A Systematic Review and Meta-Analysis of Studies Comparing Body Composition Continuous Training. Sports. 2021;9(11):1–28.

27. Pelliccia A, Sharma S, Gati S, Bäck M, Börjesson M, Caselli S, et al. 2020 ESC Guidelines on sports cardiology and exercise in patients with cardiovascular disease. Eur Heart J. 2021;42(1):17–96.

28. Abell B, Glasziou P, Hoffmann T. Reporting and Replicating Trials of Exercise-Based Cardiac Rehabilitation: Do We Know What the Researchers Actually Did? Circ Cardiovasc Qual Outcomes. 2015;8(2):187–94.

29. Treat-Jacobson D, McDermott MM, Bronas UG, Campia U, Collins TC, Criqui MH, et al. Optimal Exercise Programs for Patients with Peripheral Artery Disease: A Scientific Statement from the American Heart Association. Circulation. 2019;139(4):E10–33.

30. Slade SC, Dionne CE, Underwood M, Buchbinder R, Beck B, Bennell K, et al. Template (CERT): Modified Delphi. Phys Ther. 2016;96(10):1514–24.

31. Toval A, Solis-Urra P, Bakker EA, Sánchez-Aranda L, Fernández-Ortega J, Prieto C, et al. Exercise and brain health in patients with coronary artery disease: study protocol for the HEART-BRAIN randomized controlled trial. Front Aging Neurosci. 2024;16(August).

32. Taylor JL, Holland DJ, Spathis JG, Beetham KS, Wisløff U, Keating SE, et al. Guidelines for the delivery and monitoring of high intensity interval training in clinical populations. Prog Cardiovasc Dis [Internet]. 2019;62(2):140–6. Available from: 10.1016/j.pcad.2019.01.004

33. Bull FC, Al- SS, Biddle S, Borodulin K, Buman MP, Cardon G, et al. World Health Organization 2020 guidelines on physical activity and sedentary behaviour. 2020;1451–62.

34. Solis-Urra P, Molina-Hidalgo C, García-Rivero Y, Costa-Rodriguez C, Mora-Gonzalez J, Fernandez-Gamez B, et al. Active Gains in brain Using Exercise During Aging (AGUEDA): protocol for a randomized controlled trial. Front Hum Neurosci. 2023;17(May).

35. Fernandez-Gamez B, Solis-Urra P, Olvera-Rojas M, Molina-Hidalgo C, Fernández-Ortega J, Lara CP, et al. Resistance Exercise Program in Cognitively Normal Older Adults: CERT-Based Exercise Protocol of the AGUEDA Randomized Controlled Trial. J Nutr Heal Aging. 2023;27(10):885–93.

36. Colado JC, Garcia-Masso X, Travis Triplett N, Calatayud J, Flandez J, Behm DG, et al. Construct and concurrent validation of a new resistance intensity scale for exercise with thera-band® elastic bands. J Sport Sci Med. 2014;13(4):758–66.

37. Fernandez-Gamez B, Pulido-Muñoz Á, Olvera-Rojas M, Solis-Urra P, Corral-Pérez J, Morales JS, et al. Examining Elastic Band Properties for Exercise Prescription: AGUEDA Equations. Physiother Res Int. 2025;30(1):1–5.

38. Eston RG. Use of ratings of perceived exertion for predicting maximal work rate and prescribing exercise intensity in patients taking atenolol. Br J Sports Med. 1997;31(2):114–9.

39. Reeves GR, Gupta S, Forman DE. Evolving Role of Exercise Testing in Contemporary Cardiac Rehabilitation. 2016;309–19.

40. Eston RG, Thompson M. Use of ratings of perceived exertion for predicting maximal work rate and prescribing exercise intensity in patients taking atenolol. 1997;114–9.

41. Tibana RA, Manuel N, Sousa F De, Prestes J, Kennedy MD. Is Perceived Exertion a Useful Indicator of the Metabolic and Cardiovascular Responses to a Metabolic Conditioning Session of. (M):1–12.

42. Viana AA, Fernandes B, Alvarez C, Guimarães GV, Ciolac EG. Prescribing high-intensity interval exercise by RPE in individuals with type 2 diabetes : metabolic and hemodynamic responses. 2019;356(September 2018):348–56.

43. Hardy CJ, Rejeski WJ. Not What, But How One Feels : The Measurement of Affect During Exercise. 1989;304–17.

44. Gilgen R, Theresa A, Thomas S. RR interval signal quality of a heart rate monitor and an ECG Holter at rest and during exercise. Eur J Appl Physiol [Internet]. 2019;119(7):1525–32. Available from: 10.1007/s00421-019-04142-5

45. Montalvo S, Martinez A, Arias S, Lozano A, Gonzalez MP, Dietze-hermosa MS, et al. Commercial Smart Watches and Heart Rate Monitors : A Concurrent Validity Analysis. 2023;

46. Lo OY, Kahya M, Manor B. Powering Through Daily Activities in Older Age - Will Power Training Replace Strength Training in Later Life? JAMA Netw Open. 2022;5(5):E2211631.

47. Lucía A, Hoyos J, Santalla A, Earnest C, Chicharro JL. Tour de France versus Vuelta a España: Which is harder? Med Sci Sports Exerc. 2003;35(5):872–8.

48. Ainsworth BE, Haskell WL, Herrmann SD, Meckes N, Bassett DR, Tudor-Locke C, et al. 2011 compendium of physical activities: A second update of codes and MET values. Med Sci Sports Exerc. 2011;43(8):1575–81.

49. Weston KS, Wisløff U, Coombes JS. High-intensity interval training in patients with lifestyle-induced cardiometabolic disease: A systematic review and meta-analysis. Br J Sports Med. 2014;48(16):1227–34.

50. Zideman DA, Singletary EM, Borra V, Cassan P, Cimpoesu CD, De Buck E, et al. European Resuscitation Council Guidelines 2021: First aid. Resuscitation. 2021;161:270–90.

51. Gallagher RM, Kirkham JJ, Mason JR, Bird KA, Williamson PR, Nunn AJ, et al. Development and inter-rater reliability of the Liverpool adverse drug reaction causality assessment tool. PLoS One. 2011;6(12).

52. Shah S. Common terminology criteria for adverse events. National Cancer Institute. 2022.

53. Taylor JL, Holland DJ, Keating SE, Bonikowske AR, Coombes JS. Adherence to High-Intensity Interval Training in Cardiac Rehabilitation: A REVIEW and RECOMMENDATIONS. J Cardiopulm Rehabil Prev. 2021;41(2):61–77.

54. Mullen SP, Olson EA, Phillips SM, Szabo AN, Wójcicki TR, Mailey EL, et al. Measuring enjoyment of physical activity in older adults : invariance of the physical activity enjoyment scale (paces) across groups and time. 2011;1–9.

55. Taylor JL, Holland DJ, Keating SE, Leveritt MD, Gomersall SR, Rowlands A V., et al. Short-term and Long-term Feasibility, Safety, and Efficacy of High-Intensity Interval Training in Cardiac Rehabilitation: The FITR Heart Study Randomized Clinical Trial. JAMA Cardiol. 2020;5(12):1382–9.

56. Peixoto RP, Trombert V, Poncet A, Kizlik J, Gold G, Ehret G, et al. Feasibility and safety of high-intensity interval training for the rehabilitation of geriatric inpatients (HIITERGY) a pilot randomized study. BMC Geriatr. 2020;20(1):1–10.

57. D’Angelo MES, Pelletier LG, Reid RD, Huta V. The roles of self-efficacy and motivation in the prediction of short- and long-term adherence to exercise among patients with coronary heart disease. Heal Psychol. 2014;33(11):1344–53.

58. Bartlett JD, Close GL, Maclaren DPM, Gregson W, Drust B, Morton JP. High-intensity interval running is perceived to be more enjoyable than moderate-intensity continuous exercise: Implications for exercise adherence. J Sports Sci. 2011;29(6):547–53.

59. Rhodes RE, Kates A. Can the Affective Response to Exercise Predict Future Motives and Physical Activity Behavior? A Systematic Review of Published Evidence. Ann Behav Med. 2015;49(5):715–31.

60. Pressman SD, Petrie KJ, Sivertsen B. How strongly connected are positive affect and physical exercise? results from a large general population study of young adults. Clin Psychol Eur. 2020;2(4).

61. Hollings M, Mavros Y, Freeston J, Fiatarone Singh M. The effect of progressive resistance training on aerobic fitness and strength in adults with coronary heart disease: A systematic review and meta-analysis of randomised controlled trials. Eur J Prev Cardiol. 2017;24(12):1242–59.

62. Fan Y, Yu M, Li J, Zhang H, Liu Q, Zhao L, et al. Efficacy and Safety of Resistance Training for Coronary Heart Disease Rehabilitation: A Systematic Review of Randomized Controlled Trials. Front Cardiovasc Med. 2021;8(November):1–13.

63. Slade SC, Keating JL. Exercise prescription: A case for standardised Reporting. Br J Sports Med. 2012;46(16):1110–3.

64. Harwood AE, Russell S, Okwose NC, McGuire S, Jakovljevic DG, McGregor G. A systematic review of rehabilitation in chronic heart failure: evaluating the reporting of exercise interventions. ESC Hear Fail. 2021;8(5):3458–71.

65. Mezzani A, Hamm LF, Jones AM, McBride PE, Moholdt T, Stone JA, et al. Aerobic exercise intensity assessment and prescription in cardiac rehabilitation: A joint position statement of the European Association for Cardiovascular Prevention and Rehabilitation, the American Association of Cardiovascular and Pulmonary Rehabilitat. Eur J Prev Cardiol. 2013;20(3):442–67.

66. Bayles MP. ACSM’s Exercise Testing and Prescription. Lippincott Williams & Wilkins; 2023.

